# From Gut to Brain: A Longitudinal Assessment of Intestinal Permeability and Immune Activation Markers in Early-Stage Schizophrenia Spectrum Disorders

**DOI:** 10.64898/2026.10.16.26364494

**Authors:** Lea Jakob, Tiziana Maria Mahayri, Isis Rita Anzel Koutrouli, Kristina Kekrtová, Daniela Nováková, Tereza Totková, Filip Španiel

## Abstract

Schizophrenia spectrum disorders (SSD) have been linked to immune dysregulation and altered intestinal barrier function, although findings across studies remain inconsistent. The present study investigated circulating markers of intestinal permeability and immune activation in individuals with early-stage SSD and healthy controls and examined their associations with clinical outcomes. Serum concentrations of intestinal fatty acid-binding protein (iFABP), zonulin, soluble CD14 (sCD14), bactericidal/permeability-increasing protein (BPI), and lipopolysaccharide-binding protein (LBP) were measured in 36 patients with SSD at baseline and one-year follow-up and in 36 matched healthy controls. Linear mixed-effects models adjusted for age, sex, and body mass index (BMI) were used to evaluate group differences and longitudinal changes. No biomarker differences between patients and controls, and no longitudinal changes within patients, remained statistically significant after correction for multiple testing. Similarly, biomarker concentrations were not associated with symptom severity, global functioning, or neutrophil-to-lymphocyte ratio (NLR). Exploratory analyses identified a robust positive association between BMI and zonulin concentrations and additional associations among iFABP, LBP, and sCD14. Exploratory mixed-effects models identified a negative non-significant association between iFABP and sCD14 after adjustment for age, sex, and BMI. Overall, the findings do not support a generalized alteration of gut permeability or immune activation markers in SSD. Instead, they suggest that variation in gut-related biomarkers may be more strongly influenced by metabolic and individual factors than by diagnosis or current symptom severity.

## 1. Introduction

Inflammation, a biological response to injury or harmful stimuli aimed at defence, becomes counterproductive when it is chronic and affects the central nervous system (CNS) through neuroinflammation involving microglial activation and peripheral immune cells such as neutrophils (Butterfield et al., 2006; Müller et al., 2015; Ransohoff, 2016). The neutrophil-to-lymphocyte ratio (NLR; Chen et al., 2015), a marker of systemic inflammation and illness severity (Zhou et al., 2020), highlights the importance of neutrophils. Once they enter the CNS, neutrophils can disrupt the blood-brain barrier, exacerbate inflammation, and activate neutrophil-microglia interaction, which may lead to local damage (da Fonseca et al., 2014).

Schizophrenia spectrum disorder (SSD) diagnosis has been connected with an increase in inflammation (Müller et al., 2015), and inflammation has been highlighted as a possible step in the pathogenesis of the illness (Fond et al., 2020; Rodrigues-Amorim et al., 2018; Turner et al., 2017). The neutrophil-to-lymphocyte ratio is elevated in patients with schizophrenia (Bustan et al., 2018; Karageorgiou et al., 2019; Mazza et al., 2020; Özdin and Böke, 2019; Semiz et al., 2014), even when accounting for other metabolic parameters (Yüksel et al., 2018).

One possible mechanism may be the so-called “leaky gut” phenomenon, indicating altered intestinal permeability (Galipeau and Verdu, 2016). In this case, we note the translocation of bacterial toxins or fragments into the bloodstream, followed by inflammation and relative neutrophilia. Previous research is relatively limited in patients with schizophrenia and psychosis, only a handful of studies report elevated levels of biomarkers associated with increased intestinal permeability (Borkent et al., 2022; Gokulakrishnan et al., 2022; Severance et al., 2020; Weber et al., 2018). Additional studies have reported elevated zonulin concentrations in schizophrenia, further supporting a potential role of gut barrier dysfunction in the disorder (Barber et al., 2019; Usta et al., 2021).

Zonulin is a regulator of intestinal permeability that modulates the opening of tight junctions between epithelial cells. Increased zonulin release leads to a transient loosening of these junctions, allowing substances such as microbial components to pass from the gut into the bloodstream (Ioannou et al., 2025). Several schizophrenia studies have reported elevated circulating zonulin levels, although findings have not been consistent across patient populations (Barber et al., 2019; Usta et al., 2021), and limitations of commercial assay accuracy needs to be considered (Scheffler et al., 2018). Intestinal fatty acid binding proteins (iFABP) are small proteins (a 15-kDa intracellular protein) found in the cytosol of mature enterocytes (Galipeau and Verdu, 2016). Current schizophrenia studies (González-Blanco et al., 2024; Mukherjee et al., 2025) do not consistently support elevated iFABP; instead, iFABP appears to show weaker or distinct associations compared with bacterial translocation markers such as LBP and LPS.

Furthermore, the soluble cluster of differentiation 14 (sCD14) is a marker of monocyte activation (Borkent et al., 2022; Hoshiko et al., 2021). In association with the lipopolysaccharide-binding protein (LBP; acute inflammation phase protein, triggered by microbial translocation, originating from the liver), bacterial/permeability-increasing protein (BPI; produced by neutrophils, aimed at neutralizing microbial presence in blood, and eliminating lipopolysaccharides), and the soluble CD14 (sCD14; a marker of monocyte activation) activate signaling involved in innate immunity activation and release of proinflammatory cytokines (Borkent et al., 2022). However, the aforementioned studies have limitations that this exploratory study addresses by including two time-point measurements which enables longitudinal tracking of changes across multiple measures. As our own sample is fairly small and heterogeneous, we do not bridge the main limitations of currently available studies. Additionally, no study so far has included BPI in a human sample with SSD diagnosis.

The aims of this study are two-fold. Firstly, we aimed to investigate markers of intestinal permeability, bacterial translocation, and immune activation in individuals with early-stage schizophrenia spectrum disorders (SSD) compared with healthy controls. Specifically, we examined circulating concentrations of zonulin, intestinal fatty acid-binding protein (iFABP), lipopolysaccharide-binding protein (LBP), bacterial permeability-increasing protein (BPI), and soluble CD14 (sCD14), as well as their associations with systemic inflammation indicated by the neutrophil-to-lymphocyte ratio (NLR). Second, we aimed to examine longitudinal changes in intestinal permeability, immune activation, and clinical outcomes over a 1-year follow-up period in patients with SSD. In addition, we explored associations between biomarker concentrations and clinical measures, including symptom severity assessed using the Positive and Negative Syndrome Scale (PANSS) and global functioning assessed using the Global Assessment of Functioning (GAF) scale.

We hypothesized that patients with early-stage SSD would exhibit altered concentrations of intestinal permeability, bacterial translocation, and immune activation markers relative to healthy controls. We further hypothesized that greater intestinal permeability and immune activation would be associated with higher levels of systemic inflammation and poorer clinical status, reflected by higher PANSS scores and lower GAF scores. Finally, exploratory analyses were conducted to investigate potential biological relationships among markers of intestinal permeability, microbial translocation, and immune activation, with the aim of generating hypotheses for future mechanistic studies.

## 2. Methods

### 2.1 Participants

The participants included in this study are part of the larger ESO project (Early-stage Schizophrenia Outcome Study), a large-scale longitudinal multimodal database of patients diagnosed with SSD in the Czech Republic. As part of a complex examination, multimodal data are collected for each patient, including blood samples, MRI scans, neuropsychological functioning, stool samples, and illness severity data. The sample analyzed for this publication includes 36 first-episode SSD patients (with two-time point measures collected at a one-year interval). 36 control subjects with one measurement were included, who were tested as part of a sub-project on the gut microbiome in the SSD study.

To add, individuals with a diagnosis of SSD are included in the study following these inclusion criteria: a) undergoing first psychiatric hospitalization due to symptomatology related to SSD labeled as baseline, followed by the one-year follow-up assessment labeled as follow-up, b) during this hospitalization have received an ICD-10 diagnosis of schizophrenia (F20), acute and transient psychotic disorders (F23), or schizoaffective disorder (F25) by a psychiatrist according to ICD-10 criteria, c) < 24 months of untreated psychosis experienced before the diagnosis. All patients were hospitalized at one of the collaborating centres in Czech Republic, and were referred to the ESO study within the first month after diagnosis. Within the study, the patients were re-assessed by the psychiatrist to confirm the diagnosis and any medication history was noted.

Patients were excluded if they had: a) metabolic syndrome, b) inflammatory bowel disease, c) severe allergies (including food allergies), d) acute febrile illness at assessment, e) pregnancy or breastfeeding, f) substance or alcohol dependence, g) antibiotic use within the past 3 months, h) endocrine disorders. These criteria were primarily created for the gut microbiome project analysis, and may limit certain populations within the present sample.

Healthy participants, matched by age and sex, are included following exclusion criteria: a) lifetime history of any psychiatric disorders, b) psychotic disorders in first or second-degree relatives, c) metabolic syndrome, d) inflammatory bowel disease, e) use of chronic medication (except allergy medication and hormonal contraception), f) severe allergy, g) acute febrile illness, h) current pregnancy/ breastfeeding, i) addiction to alcohol and psychoactive substances, j) use of antibiotics in the last 3 months, k) serious endocrine diseases.

The study was approved by the Ethical Committee of the National Institute of Mental Health (approval number 203/19 and 127/17), Klecany, Czech Republic, and we adhered to the ethical standards set by national and institutional committees on human experimentation and the Declaration of Helsinki of 1975, as updated in 2008. All participants provided written informed consent.

### 2.2 Inflammatory and intestinal permeability markers

The blood samples were collected after overnight fasting. Serum was separated, aliquoted, and stored at –80 degrees Celsius with protease inhibitors until analysis. The neutrophil-to-lymphocyte ratio (NLR) was obtained from a standard blood count analysis performed by a certified clinical laboratory (AESKULAB).

We have included measures for zonulin (Human Zonulin ELISA kit, E-EL-H5560, Elabscience), intestinal fatty acid-binding proteins (IFABP/FABP2 ELISA Kit E-EL-H0159, Elabscience), soluble cluster of differentiation 14 (Human sCD14 ELISA Kit E-EL-H6149, Elabscience, dilution), lipopolysaccharide-binding protein (Human LBP ELISA Kit, E-EL-H6108, Elabscience), and bacterial/permeability-increasing protein (Human Bactericidal/permeability increasing protein (BPI) ELISA Kit, RK00987, ABclonal). All five of these kits are based on the enzyme-linked immunosorbent assay (ELISA), and include established testing protocols, intra– and inter-assay precision parameters, and sensitivity. To reduce error rates, each participant’s sample is tested in triplicate, and each kit is evaluated using a standardized procedure. The analysis of samples was carried out by trained professionals in laboratory settings, following the manufacturer’s instructions. The optical density of each well was measured using the Tecan Infinite 200 PRO universal microplate reader at 450 nm. Results were then calculated from a standard curve generated by a four-parameter logistic (4-PL) fit in R, based on the averaged triplicate values. Total protein concentration was measured using the Pierce™ Bradford Plus Protein Assay (cat. n. 23238, ThermoFisher Scientific) using the approach of Bradford (1976).

### 2.3 Data analysis approach

Statistical analyses were conducted in R, using the packages *lme4*, *lmerTest*, *emmeans*, *tidyverse*, and *broom.mixed*. Continuous biomarker variables were visually inspected and, where appropriate, log-transformed to improve normality and stabilize variance.

#### 2.3.1 Primary analyses: group differences and longitudinal changes in biomarkers

To address the primary objective of evaluating differences in gut permeability, microbial translocation, and immune activation biomarkers between patients with SSD and healthy controls, as well as longitudinal changes within patients over 1 year, linear mixed-effects models (LMMs) were fitted for each biomarker. These analyses included all available observations from healthy controls (n = 36), patients at baseline (n = 36), and patients at follow-up (n = 36). Biomarker concentrations were modeled as dependent variables, with study group (control, baseline patient, follow-up patient) included as a fixed effect. To account for within-subject dependence in the longitudinal patient measurements, subject ID was included as a random intercept. All primary models included the following covariates: age, sex, and Body mass index (biomarker ∼ group + age + sex + BMI + (1 | subject)). Estimated marginal means were calculated using the *emmeans* package, and planned contrasts were specified to compare: controls vs baseline patients, baseline vs follow-up patients, and controls vs follow-up patients.

Because biomarker concentrations were analyzed on the log scale, model coefficients were converted to percent change estimates using the transformation: Percent change = (e^A^P – 1) × 100 allowing effect sizes to be interpreted as proportional differences in biomarker concentrations between groups.

#### 2.3.2 Clinical correlates of biomarker concentrations

To address the secondary objective of evaluating whether intestinal permeability and immune activation markers were associated with clinical outcomes, Spearman correlation analyses were conducted within the patient sample using data from both assessment time points. Associations between biomarker concentrations and clinical variables were examined using Spearman correlation coefficients. Clinical measures included the Global Assessment of Functioning (GAF), Positive and Negative Syndrome Scale (PANSS) total score and subscales, and Neutrophil-to-Lymphocyte Ratio (NLR). False discovery rate (FDR) correction was applied to correlation p-values.

#### 2.3.3 Exploratory analyses of biological relationships among biomarkers

To explore potential biological relationships among intestinal permeability, enterocyte injury, microbial translocation, and immune activation markers, exploratory correlation and regression analyses were conducted within the patient sample. Spearman correlation coefficients were calculated to evaluate relationships among gut permeability markers (zonulin, iFABP), a microbial translocation marker (LBP), an immune activation marker (sCD14), and demographic and clinical variables (age, BMI, duration of untreated psychosis [DUP], and NLR). False discovery rate correction was applied to correlation p-values.

To further investigate potential mechanistic relationships, exploratory mixed-effects regression models were fitted. Specifically, the following models were examined: Enterocyte injury model: iFABP ∼ zonulin + BMI + age + sex + (1 | subject); Immune activation model: sCD14 ∼ iFABP + LBP + BMI + age + sex + (1 | subject); Full cascade model: sCD14 ∼ zonulin + iFABP + LBP + BMI + age + sex + (1 | subject)

These analyses were intended to evaluate whether markers of intestinal permeability and epithelial injury were associated with downstream markers of microbial translocation and immune activation.

#### 2.3.4 Sensitivity analyses

To evaluate the robustness of the primary findings, several sensitivity analyses were conducted. First, Winsorized analyses were performed to reduce the influence of extreme outliers. Biomarker values were Winsorized at predefined thresholds corresponding to Q1 – 1.5(IQR) and Q3 + 1.5(IQR), after which the primary models were refitted. Second, because some samples were stored for an extended period, additional mixed-effects models were estimated incorporating Bradford protein concentration as a covariate to account for potential variation attributable to total protein concentration. Third, complementary non-parametric analyses were conducted using Wilcoxon rank-sum tests for cross-sectional comparisons and Wilcoxon signed-rank tests for longitudinal comparisons. Additional patient-only sensitivity analyses examined whether longitudinal biomarker trajectories were influenced by chlorpromazine-equivalent antipsychotic exposure or duration of untreated psychosis (DUP).

#### 2.3.5 Multiple testing and data visualization

To account for multiple comparisons across biomarkers and planned contrasts, false discovery rate (FDR) correction was applied using the Benjamini–Hochberg procedure. Biomarker distributions across groups were visualized using violin plots combined with boxplots and jittered observations. Longitudinal patient trajectories were visualized using spaghetti plots, and relationships between biomarkers were visualized using scatter plots with fitted regression lines and confidence intervals.

## 3. Results

### 3.1 Participant characteristics

Participant demographics and clinical characteristics across three groups: healthy controls (n = 36), patients at baseline (n = 36), and the same group of patients at a later follow-up (n = 36) are shown in Table 1. Age was well-matched across groups, with mean ages of 28.0 (SD = 5.36), 27.04 (SD = 8.06), and 28.1 (SD = 8.04). Overall, we included 16 women in the control group (and 16 in the patient group), and 20 men in the control group (matched to 20 men in the patient group).

**Table 1.**
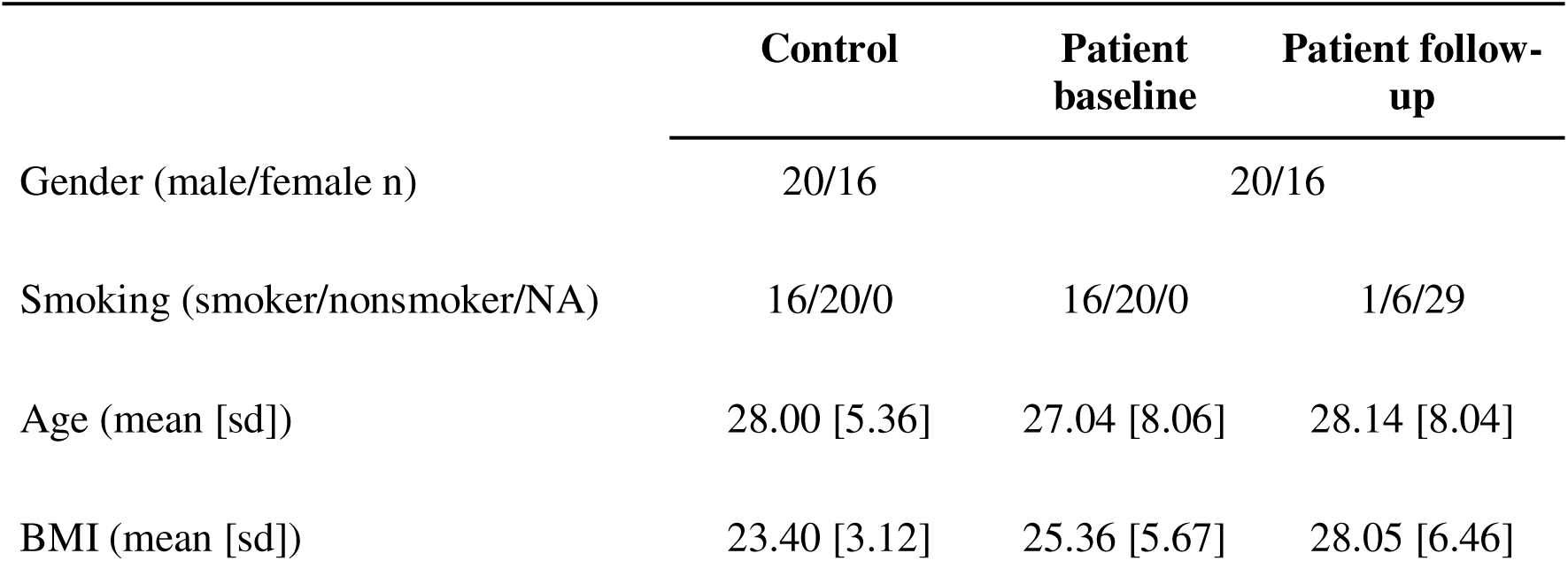
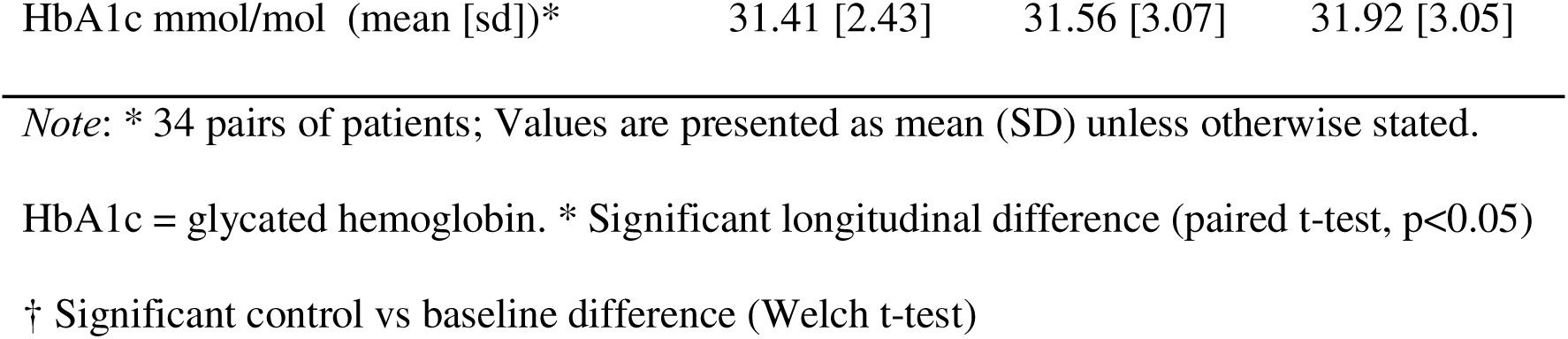
Descriptive data for the patient (n = 36) and control sample (n = 36)

Body mass index (BMI) increased across time in patients, from a baseline mean of 25.36 (SD = 5.67) to 28.05 (SD = 6.46) at follow-up, while controls averaged 23.4 (SD = 3.12), while the glycated hemoglobin did not differ across groups.

Welch’s t-test for age between patient baseline and controls was not statistically significant (W = 0.598, p = 0.552). BMI comparison of the patient baseline timepoint and control group, there was no statistically significant difference (Welch’s W = –1.819, p = 0.074), and the change between two patient visits was also not significant (paired t-test t = – 5.260, p < 0.001). Lastly, the glycated hemoglobin values were not significantly different between the baseline patient timepoint and control group (Welch’s W = –0.225, p = 0.823), and the difference within patients did not significantly change based on a paired t-test (t = – 1.200, p = 0.239).

#### Patient sample

Diagnoses according to ICD codes revealed that the majority of patients were diagnosed with schizophrenia (F20.0; 58.3%), with smaller proportions assigned to other related psychotic disorders (e.g., F23.1; 23.6%, F23.0; 11.1%, F25.2; 2.8%, and one patient each with diagnoses F20.5, F20.8, F25.0), making the sample heterogenous, while still all patients belong in the Schizophrenia Spectrum Disorder diagnosis cluster. Psychopathology assessments showed improvement over time, with statistically significant change (see Table 2). The PANSS total score decreased from 56.97 (SD = 12.93) at baseline to 49.03 (SD = 13.54) at follow-up. General functioning, measured using the GAF, increased from a mean of 64.75 (SD = 16.25) to 74.44 (SD = 13.00), indicating an improvement in patients’ overall functioning. In the one year interval, the patients had statistically significantly reduced chlorpromazine equivalent load of medication (Wilcoxon’s V = 497, p < 0.001). Duration of untreated psychosis (DUP) was defined as the interval between the patient’s first psychotic symptoms and initiation of antipsychotic treatment. Although participants were recruited during their first psychiatric hospitalization, baseline research assessments did not necessarily occur on the first day of treatment initiation. In this sample, DUP was highly variable (M = 3.30 months, SD = 5.14), as outlined in Table 2, indicating on average the time between self-perceived first symptoms and the established diagnosis with treatment initiation was around 3 months. Consequently, some patients had already received antipsychotic treatment before baseline blood sampling (e.g., in a different hospital), explaining the non-zero treatment duration reported in Table 2.

**Table 2.**
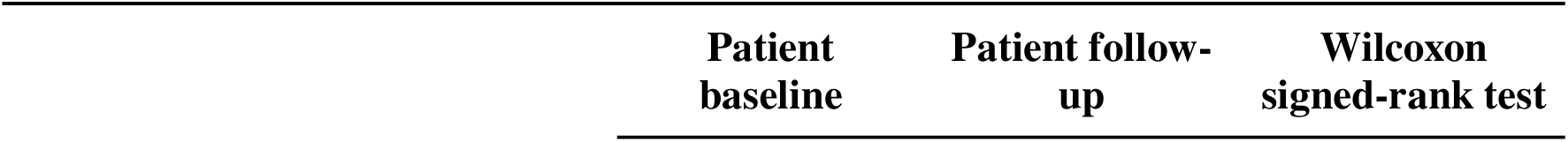
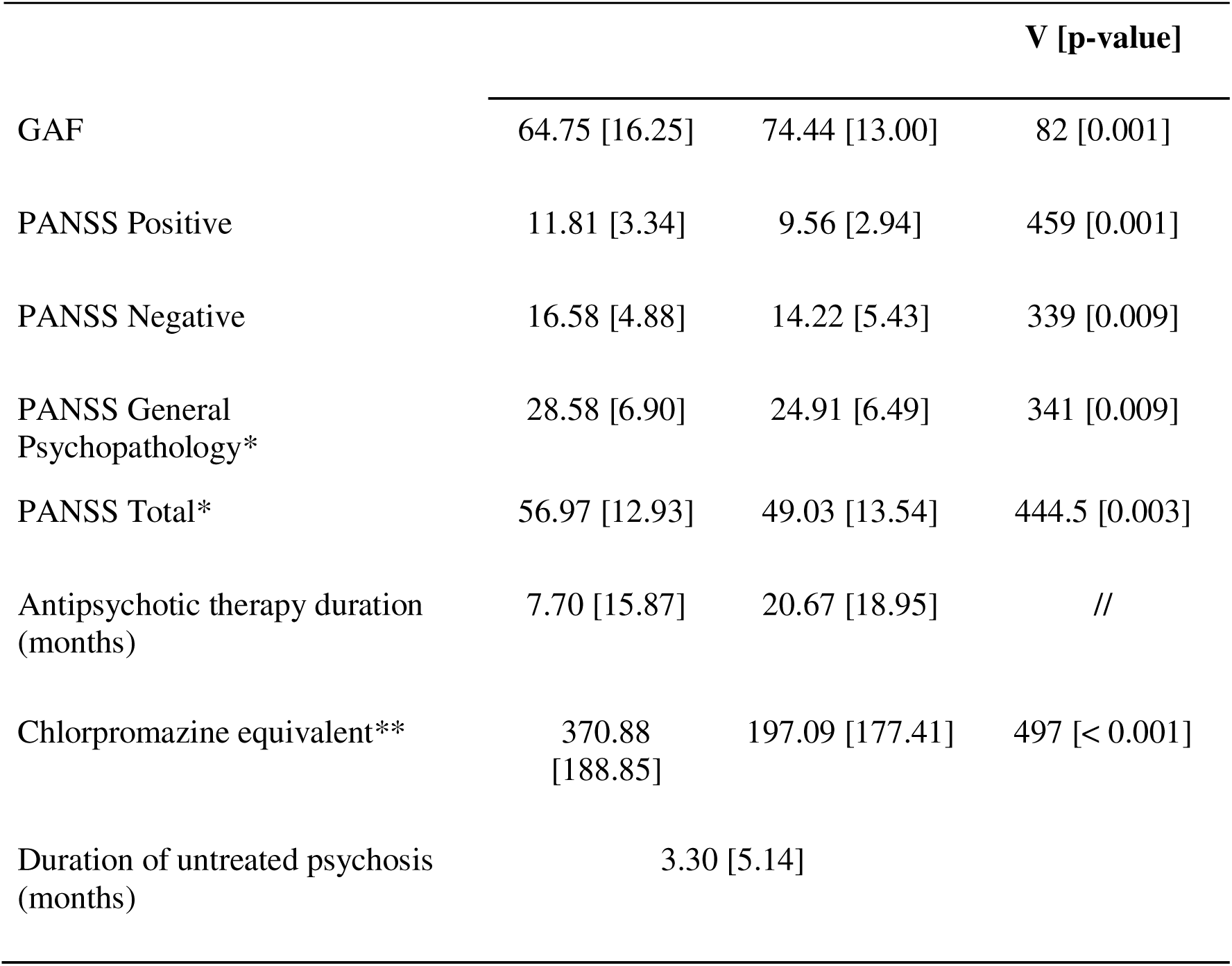
Descriptive data for the patient (n = 36) sample clinical characteristics. *Note*: Values are presented as mean (SD). Longitudinal differences were evaluated using Wilcoxon signed-rank tests. P-values represent exploratory within-patient comparisons based on Wilcoxon signed-rank tests and are provided for descriptive purposes only. Primary inferential analyses are reported in Table 4. GAF = Global Assessment of Functioning; PANSS = Positive and Negative Syndrome Scale; *34 paired observations available; **35 paired observations available.

We conducted enzyme-linked immunosorbent assays (ELISAs) to assess serum biomarkers of intestinal permeability in patients with SSD and healthy controls. The main descriptive statistics for the immune parameters and the intestinal permeability measures are shown in Table 3, Figure 1(A) and Table S1.

**Figure 1.**
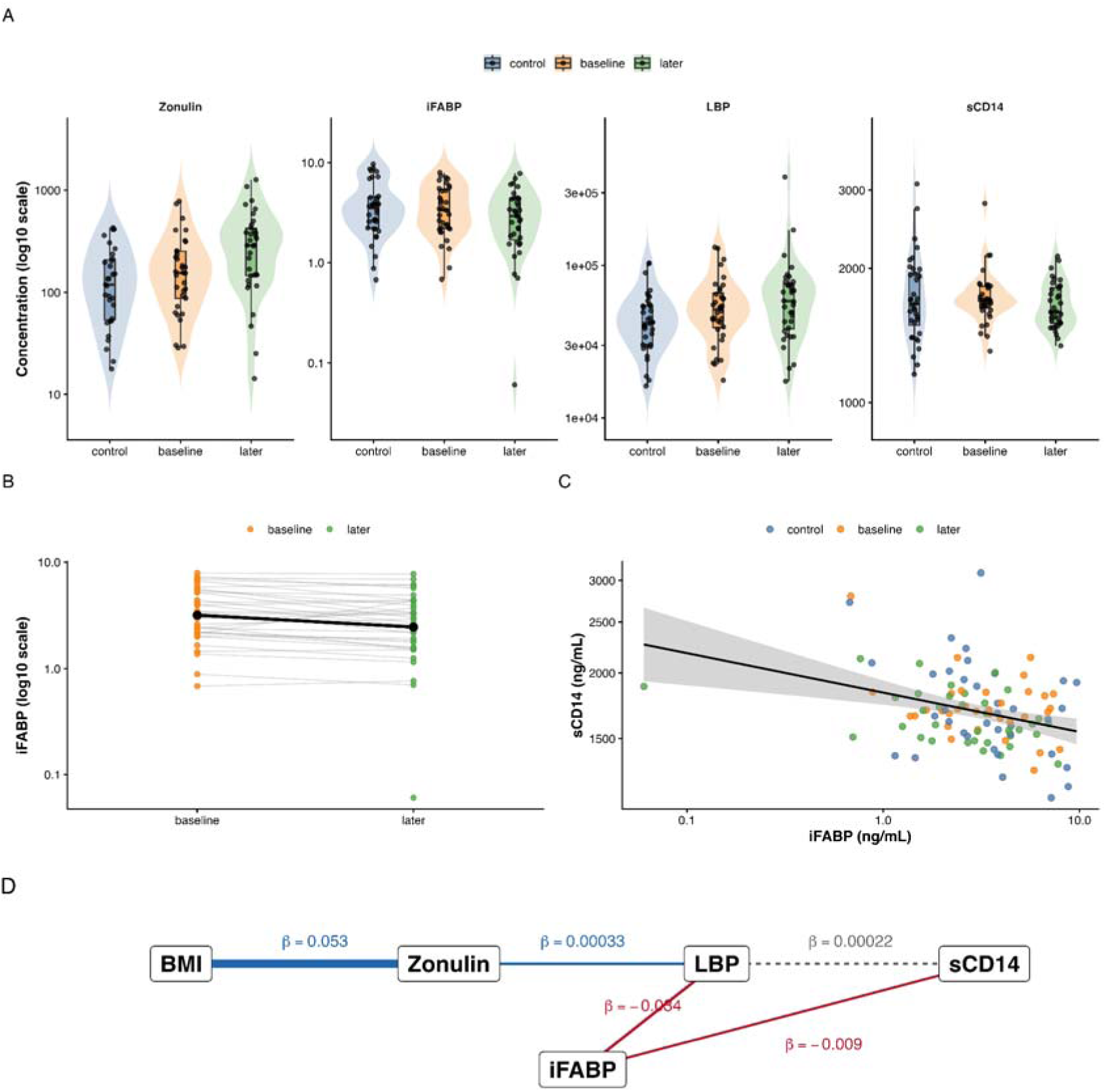
Gut barrier dysfunction and immune activation in schizophrenia. (A) Distribution of selected biomarkers across study groups (controls, baseline patients, and follow-up patients) shown on a log scale. (B) Individual patient trajectories for iFABP concentrations between baseline and follow-up. Grey lines connect repeated measurements from the same participant, black dots and line represent group means. (C) Association between iFABP and sCD14 across all available observations, after adjustment for covariates. (D) Exploratory pathway model summarizing significant associations identified in mixed-effects analyses. Positive associations are shown in blue and negative associations in red.

**Table 3.**
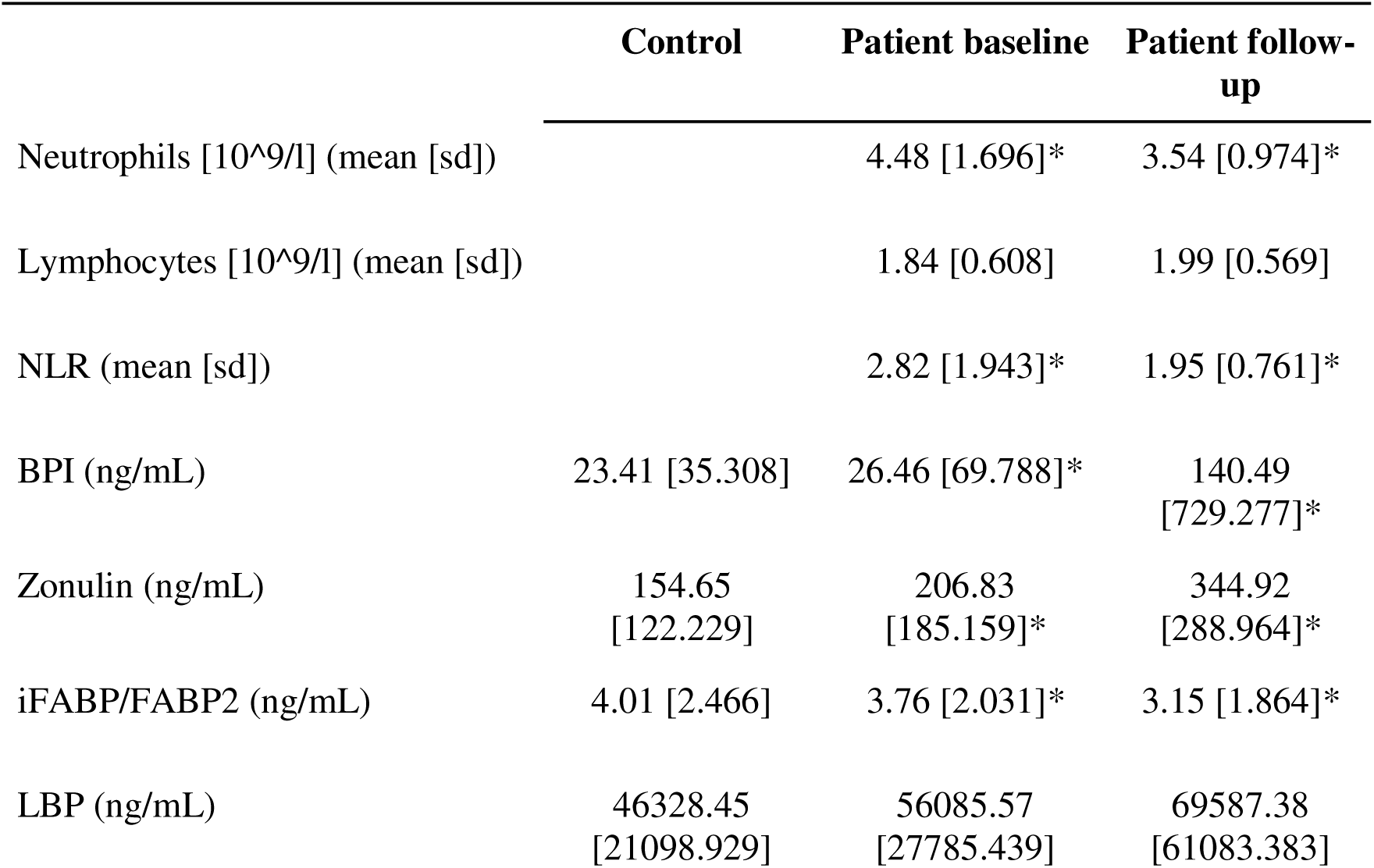
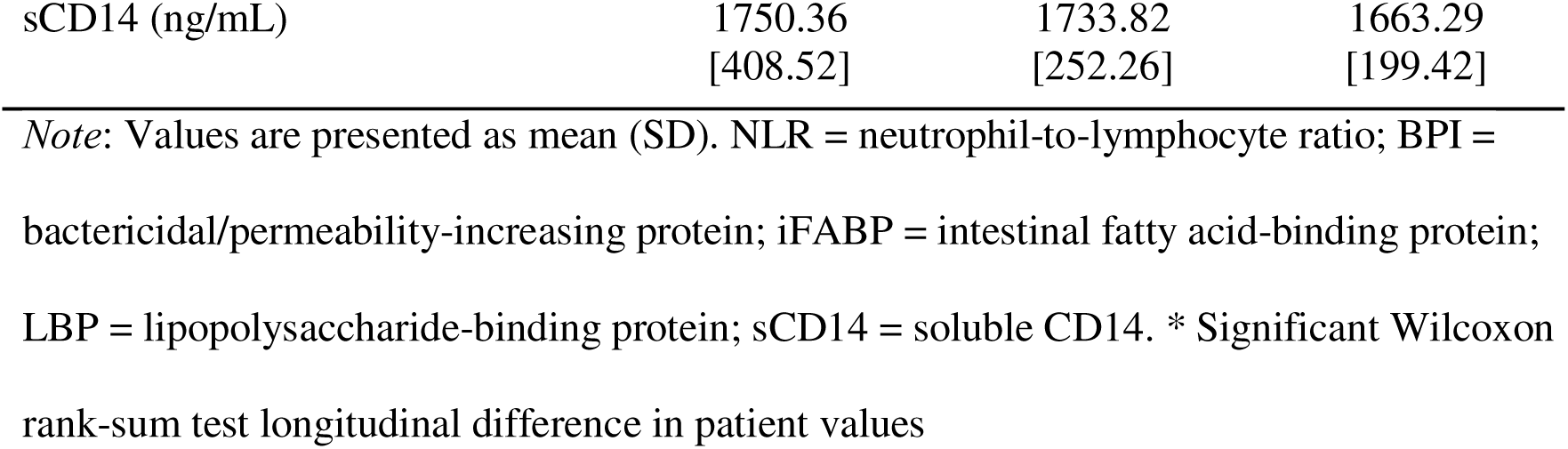
Descriptive data of intestinal permeability parameters for the patient (n = 36) and control sample (n = 36)

|  | Control | Patient baseline | Patient follow-up |
| --- | --- | --- | --- |
| Neutrophils [$10^9/l$] (mean [sd]) | | 4.48 [1.696]* | 3.54 [0.974]* |
| Lymphocytes [$10^9/l$] (mean [sd]) | | 1.84 [0.608] | 1.99 [0.569] |
| NLR (mean [sd]) |  | 2.82 [1.943]* | 1.95 [0.761]* |
| BPI (ng/mL) | 23.41 [35.308] | 26.46 [69.788]* | 140.49 [729.277]* |
| Zonulin (ng/mL) | 154.65 [122.229] | 206.83 [185.159]* | 344.92 [288.964]* |
| iFABP/FABP2 (ng/mL) | 4.01 [2.466] | 3.76 [2.031]* | 3.15 [1.864]* |
| LBP (ng/mL) | 46328.45 [21098.929] | 56085.57 [27785.439] | 69587.38 [61083.383] |
| sCD14 (ng/mL) | 1750.36<br>[408.52] | 1733.82<br>[252.26] | 1663.29<br>[199.42] |

*Note:* Values are presented as mean (SD). NLR = neutrophil-to-lymphocyte ratio; BPI = bactericidal/permeability-increasing protein; iFABP = intestinal fatty acid-binding protein; LBP = lipopolysaccharide-binding protein; sCD14 = soluble CD14. \* Significant Wilcoxon rank-sum test longitudinal difference in patient values

Considering the comparison of the control values and the baseline values for the patient sample, there were no statistically significant differences in intestinal permeability markers based on the Wilcoxon rank-sum test (all p-values were above the 0.05 threshold). Comparing patient values at baseline to follow-up, the Wilcoxon rank-sum test showed statistically significant differences in Neutrophils (V = 356, p = 0.035), NLR (V = 392, p = 0.047), BPI (V = 186, p = 0.035), zonulin (V = 60, p < 0.001), iFABP/iFABP2 (V = 461, p = 0.005). All other parameters were not significantly different. A more complex analysis is shown in the next section.

Higher body mass index (BMI) was associated with increased zonulin levels, and higher zonulin levels were associated with increased LBP concentrations. In contrast, higher iFABP levels were associated with lower LBP and lower sCD14 concentrations. A positive association between LBP and sCD14 was observed but did not survive multiple-testing correction (dashed grey line). The pathway diagram is intended as a visual summary of exploratory relationships and does not imply causal directionality.

### 3.2 Objective 1: Group differences and longitudinal changes in gut permeability biomarkers

Linear mixed-effects models adjusted for age, sex, and BMI revealed no statistically significant differences in biomarker concentrations after false-discovery-rate (FDR) correction (Table 4). Among the planned contrasts, the strongest longitudinal trend was observed for iFABP, which decreased between baseline and follow-up (β = –0.173, p = 0.005, q = 0.076), corresponding to an estimated 15.9% reduction in circulating concentrations. Individual patient trajectories for iFABP are illustrated in Figure 1B. A nominal difference was also observed for LBP, with higher concentrations in follow-up patients compared with controls (β = 0.323, p = 0.014, q = 0.102). However, neither association remained statistically significant after correction for multiple testing.

**Table 4.**
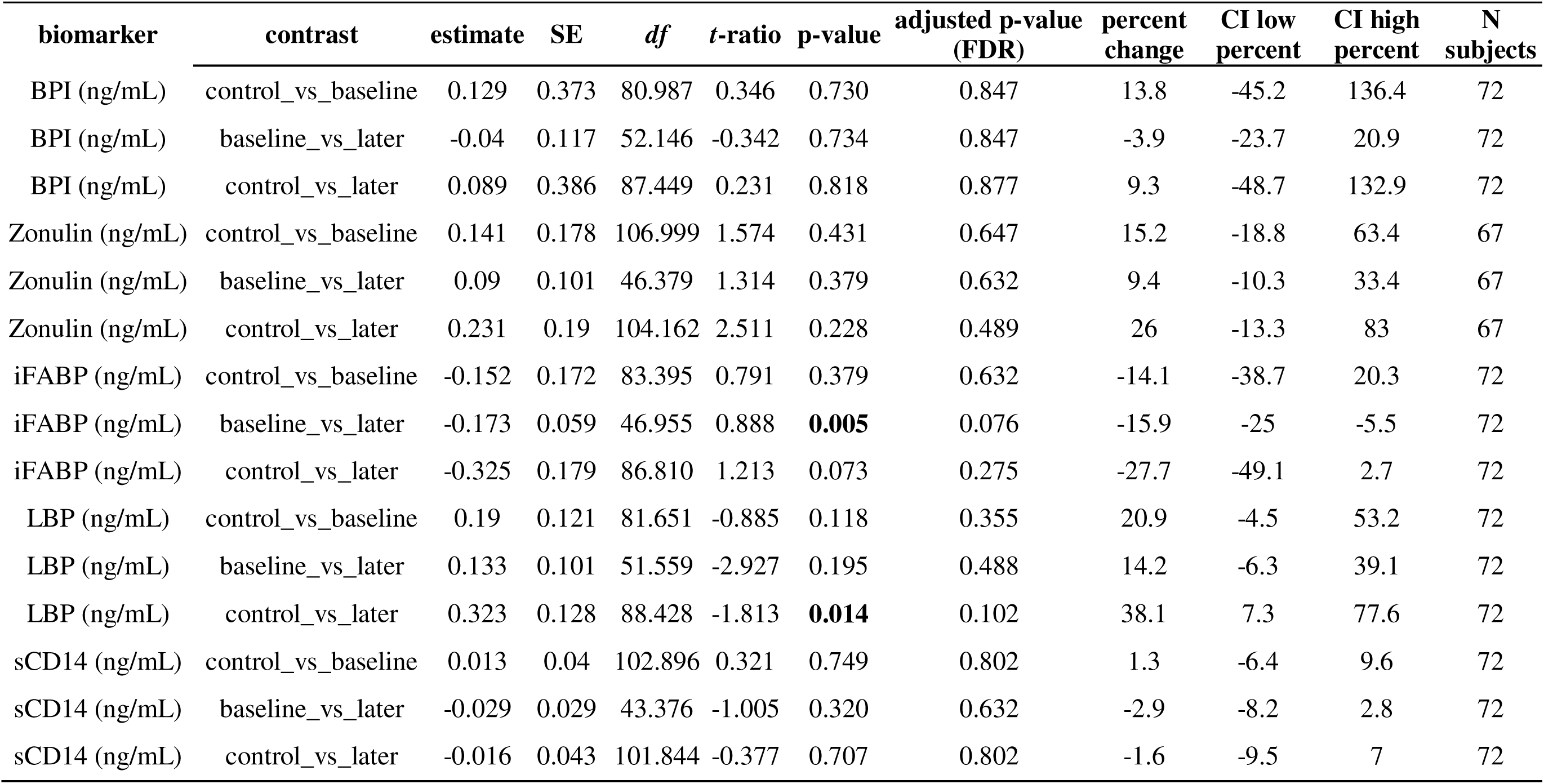
Primary mixed model results: Biomarker group-level contrasts with percent change indicators.

| biomarker | contrast | estimate | SE | df | t-ratio | p-value | adjusted p-value<br>(FDR) | percent<br>change | CI low<br>percent | CI high<br>percent | N<br>subjects |
| --- | --- | --- | --- | --- | --- | --- | --- | --- | --- | --- | --- |
| BPI (ng/mL) | control_vs_baseline | 0.129 | 0.373 | 80.987 | 0.346 | 0.730 | 0.847 | 13.8 | -45.2 | 136.4 | 72 |
| BPI (ng/mL) | baseline_vs_later | -0.04 | 0.117 | 52.146 | -0.342 | 0.734 | 0.847 | -3.9 | -23.7 | 20.9 | 72 |
| BPI (ng/mL) | control_vs_later | 0.089 | 0.386 | 87.449 | 0.231 | 0.818 | 0.877 | 9.3 | -48.7 | 132.9 | 72 |
| Zonulin (ng/mL) | control_vs_baseline | 0.141 | 0.178 | 106.999 | 1.574 | 0.431 | 0.647 | 15.2 | -18.8 | 63.4 | 67 |
| Zonulin (ng/mL) | baseline_vs_later | 0.09 | 0.101 | 46.379 | 1.314 | 0.379 | 0.632 | 9.4 | -10.3 | 33.4 | 67 |
| Zonulin (ng/mL) | control_vs_later | 0.231 | 0.19 | 104.162 | 2.511 | 0.228 | 0.489 | 26 | -13.3 | 83 | 67 |
| iFABP (ng/mL) | control_vs_baseline | -0.152 | 0.172 | 83.395 | 0.791 | 0.379 | 0.632 | -14.1 | -38.7 | 20.3 | 72 |
| iFABP (ng/mL) | baseline_vs_later | -0.173 | 0.059 | 46.955 | 0.888 | **0.005** | 0.076 | -15.9 | -25 | -5.5 | 72 |
| iFABP (ng/mL) | control_vs_later | -0.325 | 0.179 | 86.810 | 1.213 | 0.073 | 0.275 | -27.7 | -49.1 | 2.7 | 72 |
| LBP (ng/mL) | control_vs_baseline | 0.19 | 0.121 | 81.651 | -0.885 | 0.118 | 0.355 | 20.9 | -4.5 | 53.2 | 72 |
| LBP (ng/mL) | baseline_vs_later | 0.133 | 0.101 | 51.559 | -2.927 | 0.195 | 0.488 | 14.2 | -6.3 | 39.1 | 72 |
| LBP (ng/mL) | control_vs_later | 0.323 | 0.128 | 88.428 | -1.813 | **0.014** | 0.102 | 38.1 | 7.3 | 77.6 | 72 |
| sCD14 (ng/mL) | control_vs_baseline | 0.013 | 0.04 | 102.896 | 0.321 | 0.749 | 0.802 | 1.3 | -6.4 | 9.6 | 72 |
| sCD14 (ng/mL) | baseline_vs_later | -0.029 | 0.029 | 43.376 | -1.005 | 0.320 | 0.632 | -2.9 | -8.2 | 2.8 | 72 |
| sCD14 (ng/mL) | control_vs_later | -0.016 | 0.043 | 101.844 | -0.377 | 0.707 | 0.802 | -1.6 | -9.5 | 7 | 72 |

Sensitivity analyses using Winsorized data, Bradford-adjusted models, and non-parametric testing yielded comparable results (Tables S2a–S4 and Figure S1). In models examining covariates (Table S5), higher BMI was associated with higher zonulin concentrations (β = 0.111, p < 0.001), whereas male sex was associated with lower zonulin concentrations (β = –0.952, p < 0.001) and higher iFABP concentrations (β = 0.548, p = 0.002). No consistent associations were observed for age. Additional sensitivity analyses adjusting for chlorpromazine-equivalent antipsychotic exposure (Table S6) and duration of untreated psychosis (DUP; Table S7) yielded a comparable pattern of findings. No biomarker group differences remained statistically significant after FDR correction. In the DUP-adjusted models, the longitudinal decrease in iFABP remained significant (β = –0.206, p = 0.002, q = 0.010), whereas no corresponding effects were observed for the remaining biomarkers.

### 3.3 Objective 2: Associations between biomarkers and clinical outcomes

Associations between biomarker concentrations and clinical measures were evaluated within the patient sample using Spearman correlation analyses (Table S8). No significant associations were observed between biomarker concentrations and GAF scores, PANSS total scores, or PANSS subscale scores after correction for multiple testing. Thus, the hypothesis that greater intestinal permeability or immune activation would be associated with poorer clinical outcomes was not supported.

### 3.4 Exploratory analyses of biological relationships among biomarkers

Correlation analyses revealed one association that survived FDR correction: higher BMI was associated with higher zonulin concentrations (ρ = 0.65, q < 0.001; Table S9). No significant associations were observed between NLR and any gut permeability or immune activation marker.

Exploratory mixed-effects regression analyses suggested two partially distinct biological processes (Figure 1D). First, higher BMI was associated with increased zonulin concentrations, which in turn were associated with higher LBP concentrations. Second, higher iFABP concentrations were associated with lower LBP and lower sCD14 concentrations. In the exploratory immune activation model, iFABP was negatively associated with sCD14 after adjustment for age, sex, and BMI (β = –0.062, p = 0.010). This relationship is illustrated in Figure 1C.

Together, these findings do not support a simple sequential pathway from intestinal injury to systemic immune activation but instead suggest that markers of epithelial injury and microbial translocation may capture distinct aspects of gut–immune dysfunction in SSD.

## 4. Discussion

The present study investigated circulating markers of intestinal permeability and immune activation in individuals with schizophrenia spectrum disorders (SSD) using both cross-sectional and longitudinal analyses. Contrary to our primary hypothesis, we found no robust differences between patients and healthy controls and no biomarker changes that remained significant after correction for multiple testing. Similarly, biomarkers of intestinal permeability and immune activation were not associated with symptom severity or global functioning. Exploratory analyses nevertheless identified several relationships among gut-related biomarkers and metabolic variables, particularly between BMI and zonulin, and between iFABP, LBP, and sCD14. Together, these findings suggest that gut-related biological variation in SSD may be more strongly influenced by metabolic and individual factors than by diagnostic status or current symptom severity.

### 4.1 Objective 1: Group differences and longitudinal changes in gut permeability biomarkers

The primary objective of this study was to determine whether markers of intestinal permeability and immune activation differ between individuals with SSD and healthy controls, and whether these markers change longitudinally during treatment. Overall, the findings did not support this hypothesis. None of the examined biomarkers showed statistically significant group differences after correction for multiple testing, and no longitudinal changes remained significant after FDR adjustment.

Gut permeability markers (zonulin, sCD14, LBP) have been reported to be elevated across psychiatric disorders and correlate with “sickness behavior” (Safadi et al., 2022). However, studies focusing on patients with a diagnosis of SSD have reported inconsistent changes. For example, iFABP has been reported as lower in acute antipsychotic-free schizophrenia compared with controls in the study by Mukherjee et al. (2025), whereas Jensen et al. (2023) reported higher plasma iFABP and LBP levels in both schizophrenia and affective disorders, and González-Blanco et al. (2024) found only weak associations of iFABP and LPS with symptom dimensions that disappeared after adjustment for age and BMI.

One exploratory finding was an association between iFABP and sCD14 that remained significant after adjustment for BMI, age, and sex. Because iFABP is commonly interpreted as a marker of enterocyte injury and sCD14 as a marker of innate immune activation, this observation may indicate a relationship between intestinal epithelial integrity and immune signaling. However, the association was identified in exploratory analyses and should be interpreted cautiously. Rather than demonstrating a causal pathway, these findings suggest that enterocyte injury and immune activation may represent interconnected biological processes that warrant further investigation in larger longitudinal cohorts. Mukherjee et al. (2025) also reported that LBP correlated strongly with neutrophils in acute schizophrenia, suggesting an interface between gut-derived inflammatory signals and peripheral immune activation. However, because our data are observational, this association should still be interpreted cautiously as evidence of co-occurring biological processes rather than proof of causality.

Similarly, the outcomes related to sCD14 and LBP in this population are mixed. Elevated sCD14 has been reported in schizophrenia by Severance et al. (2013), who found that sCD14 seropositivity was associated with a 3.1-fold increased odds of schizophrenia. When it comes to the stage of illness, Dzikowski et al. (2020) reported significantly higher sCD14, especially in chronic schizophrenia, while Weber et al. (2018) found elevated sCD14 before schizophrenia diagnosis, suggesting that innate immune activation may predate illness onset. LBP findings have been less consistent: Severance et al. (2013) did not find matched case–control differences in LBP, Weber et al. (2018) reported unchanged or marginally decreased LBP before diagnosis, Brokent et al. (2020) and Scheurink et al. (2023) found no group differences in either LBP or sCD14 between schizophrenia spectrum disorder patients and controls. Furthermore, Mørch et al. (2019) reported that sCD14 tended to be lower in psychotic disorders after adjustment for confounders. However, this was not specific to schizophrenia status as their investigation focused on individuals with a psychotic episode (either as part of the diagnosis of schizophrenia, major depressive disorder, or compared with order). Compared with the mixed results of these studies, the lack of robust case–control differences in our study after multiple-testing correction supports the view that these biomarkers are unlikely to serve as stable trait markers of schizophrenia across cohorts and illness stages.

Although no longitudinal biomarker changes survived correction for multiple testing, iFABP showed the strongest longitudinal trend, with lower concentrations observed at follow-up compared with baseline. This finding should be interpreted cautiously given the absence of FDR-corrected significance, but it may indicate that some aspects of intestinal epithelial injury are more dynamic than other components of the gut–immune axis. In contrast, no comparable longitudinal changes were observed for sCD14, suggesting that markers of immune activation may follow different trajectories over time. This interpretation is broadly compatible with studies showing abnormal iFABP or sCD14 levels in acute or early-phase samples, including Mukherjee et al. (2025), Dzikowski et al. (2020), and Weber et al. (2018), and helps explain why cross-sectional studies at different illness stages have produced conflicting results. At the same time, our observation that LBP was higher in follow-up patients compared with controls suggests that endotoxin-related exposure may persist or emerge later, even when markers of epithelial injury decline. This may help explain why some studies detect elevated LBP in schizophrenia (Jensen et al., 2023; Mukherjee et al., 2025), whereas others do not (Scheurink et al., 2023; Severance et al., 2013; Weber et al., 2018). One possibility is that LBP reflects a more chronic or metabolism-driven process, whereas iFABP and sCD14 may capture more dynamic changes related to acute epithelial stress and immune activation.

The BMI findings we presented are also consistent with prior studies suggesting that metabolic factors contribute importantly to variation in gut-related biomarkers, especially impacted in SSD. González-Blanco et al. (2024) reported associations between BMI and both LBP and LPS, while Severance et al. (2013) found that quantitative LBP levels correlated with BMI in schizophrenia. In our data, the association between BMI and zonulin emerged consistently across both mixed-effects and correlation analyses, making it one of the most robust findings of the study, whereas no consistent associations were observed between BMI and the other gut-related biomarkers. This interpretation is consistent with a metabolic endotoxemia framework, in which higher adiposity contributes to barrier dysfunction. However, age showed no clear association with biomarker levels in our sample, which is broadly consistent with González-Blanco et al. (2024), who did not find clear effects of age on LBP or iFABP in their schizophrenia sample.

It is important to mention the possible role of antipsychotic and general psychiatric treatment on the related metabolic changes of the present study sample. Although participants were recruited close to illness onset, they were not medication-naïve, and antipsychotic treatment duration (but not the chlorpromazine exposure equivalent) increased substantially during follow-up. Antipsychotic medications are known to influence body weight, metabolic function, inflammatory pathways, and gut microbial composition. Consequently, longitudinal biomarker changes may reflect treatment-related physiological adaptations rather than illness progression alone. Importantly, sensitivity analyses adjusting for chlorpromazine-equivalent exposure did not materially alter the overall pattern of results, suggesting that medication effects are unlikely to fully explain the observed findings. Nevertheless, future studies beginning prior to treatment initiation would be valuable for disentangling illness-related and medication-related effects on the gut–immune axis.

### 4.2 Objective 2: Clinical correlates

The absence of significant associations between gut biomarkers and symptom severity in our sample also fits with previous literature. González-Blanco et al. (2024) reported weak positive correlations of iFABP and LPS with some symptom measures, particularly negative symptoms. Still, these associations were no longer significant after adjustment for age and BMI. Our null findings for PANSS, GAF, and NLR therefore support the view that gut permeability and immune activation markers may not map directly onto current clinical severity, or that these relationships are subtle and highly confounded by metabolic and demographic factors. Similarly, our finding that longer duration of untreated psychosis was not clearly associated with worse biomarker levels suggests that these biological processes may not simply relate to untreated illness.

Finally, the sex differences in our sample may represent an important source of heterogeneity across studies. Severance et al. (2013) reported higher sCD14 and LBP levels in women, whereas in our sample, men showed higher iFABP and lower zonulin concentrations. Although these findings are not directly aligned, together they suggest that sex-related differences in gut barrier physiology, immune activation, or body composition may contribute substantially to variability in biomarker profiles across cohorts. This may partly explain why the literature remains inconsistent and supports the need for sex-stratified analyses in future work. Overall, our results fit best with a model in which gut barrier dysfunction and immune activation in SSD are not uniform disease traits, but dynamic and heterogeneous processes shaped by illness stage, metabolic status, and sex (Dzikowski et al., 2020; González-Blanco et al., 2024; Jensen et al., 2023; Mørch et al., 2019; Mukherjee et al., 2025; Scheurink et al., 2023; Severance et al., 2013; Weber et al., 2018).

### 4.3 Alternative model

Rather than a disease-specific gut permeability model, our findings may support a heterogeneity model of gut–immune dysfunction in SSD given the predominantly negative findings. The previously cited literature implicitly assumes a generalized disease model in which increased intestinal permeability leads to microbial translocation, systemic immune activation, and lastly leads to psychotic symptom emergence or worsening. If we are to consider these models to be true, we would expect to find relatively consistent elevations in gut permeability markers among patients and meaningful associations with symptom severity. However, none of these predictions were supported by our data, even with their limitations.

Instead, our findings may be more consistent with a metabolic–gut–immune heterogeneity model. In this framework, we consider gut permeability and immune activation not to be viewed as universal features of SSD but rather as biological processes that vary across individuals and may be influenced by metabolic status, sex, treatment exposure, lifestyle factors, and other sources of heterogeneity. Several observations based on our data support this interpretation. First, no biomarker demonstrated a robust case–control differences after correction for multiple testing. Second, gut-related biomarkers were not associated with symptom severity or functional outcomes. Third, BMI emerged as one of the strongest correlates of biomarker variation, particularly for zonulin concentrations. Finally, exploratory analyses suggested relationships among biomarkers that appeared to be partly independent of diagnostic status.

This interpretation is also compatible with growing evidence that schizophrenia is biologically heterogeneous and may comprise multiple partially overlapping pathophysiological processes rather than a single disease mechanism (Alnæs et al., 2019; Clementz et al., 2016; Duncan et al., 2025; Ivleva et al., 2017). Within such a framework, gut–immune dysregulation may characterize only a subset of patients, potentially those with greater metabolic burden, inflammatory activation, altered microbiome composition, or specific environmental exposures. Consequently, future studies may benefit from stratifying patients according to metabolic and inflammatory phenotypes rather than treating SSD as a biologically homogeneous group.

Although speculative, this model may help explain why previous studies have reported inconsistent findings regarding iFABP, LBP, sCD14, and zonulin across different schizophrenia cohorts. Differences in illness stage, medication exposure, metabolic status, and recruitment strategies may influence the proportion of individuals expressing gut– immune abnormalities, resulting in variable findings across studies. Future longitudinal investigations incorporating direct measures of intestinal permeability, microbiome composition, metabolic dysfunction, and immune activation will be needed to evaluate this hypothesis more directly.

### 4.4 Strengths and Limitations of the Present Study

Strengths of this study include its longitudinal design, the inclusion of a matched control group, the assessment of multiple complementary biomarkers not previously reported in the same sample, and the use of statistical models that adjust for key covariates that may affect interpretation.

Nevertheless, several limitations should be considered when interpreting these findings. First, although the study included longitudinal patient data, the sample size was small, particularly for patient-only analyses. This limits statistical power to detect group differences or longitudinal changes after correction for multiple comparisons. Some analytes had missing values because concentrations exceeded the detection limit or were under the limit of detection, while for some clinical and demographic measures, patients did not provide sufficient information to be used for analyses, such as the smoking status as follow-up. Second, biomarker measurements are indirect indicators of gut barrier function and microbial translocation, and direct measures of intestinal permeability or microbiome composition were unavailable. It is important to mention that while zonulin is commonly used as a marker of intestinal permeability; however, findings should be interpreted cautiously because widely used commercial assays may not detect zonulin/pre-haptoglobin-2 specifically, instead detecting other proteins such as properdin, haptoglobin, or complement C3 (Scheffler et al., 2018). Third, the diagnostic data show high heterogeneity among patients, with multiple psychotic disorder codes (F20.0, F23.1, F25.0), which could reflect underlying differences in pathophysiology. Without stratifying by diagnosis or inflammatory phenotype, interpretation remains broad and exploratory. Additionally, medication status and type (due to heterogeneity in types, combinations, and dosages), diet, and lifestyle factors, which may influence gut permeability and immune activation, were not fully accounted for in the analyses while previous studies point to the fact that these variables are relevant to gut microbiome as well as intestinal permeability. Lastly, our recruitment strategy limited the sample to individuals without metabolic syndrome, inflammatory bowel disease, allergies, or recent antibiotic use. This additionally limits any conclusions we may make, as the population of patients with these health states are common, and therefore artificially excluded from our model.

These findings do not provide strong support for a generalized gut permeability abnormality in SSD but suggest that gut-related biomarkers may be influenced by metabolic status and individual biological heterogeneity. Additionally, our model does not confirm that this process is specific to patients, implying the possibility of a general biological process.

## 5. Conclusion

In this longitudinal study of individuals with schizophrenia spectrum disorders (SSD), we did not identify robust differences in circulating markers of intestinal permeability and immune activation compared with healthy controls after correction for multiple comparisons. Similarly, biomarker concentrations were not associated with symptom severity or functional outcomes, providing no support for the hypothesis that greater gut permeability or immune activation is directly related to clinical presentation in SSD. Exploratory analyses nevertheless suggested relationships among gut-related biomarkers and metabolic factors. In particular, higher BMI was consistently associated with higher zonulin concentrations, and additional associations were observed among iFABP, LBP, and sCD14. These findings suggest that gut-related biological variation may be influenced more strongly by metabolic and individual characteristics than by diagnostic status alone.

Taken together, our results do not support a generalized model of gut barrier dysfunction as a stable feature of SSD. Instead, they are more consistent with a heterogeneous metabolic–gut–immune framework, in which gut permeability and immune activation may characterize specific patient subgroups rather than the disorder as a whole. Future studies with larger samples, direct measures of intestinal permeability and microbiome composition, and more detailed characterization of metabolic and treatment-related factors will be needed to determine whether distinct gut–immune phenotypes exist within SSD and whether they have clinical relevance.

## Data Availability

All data produced in the present study are available upon reasonable request to the authors

## Acknowledgements

This project has been supported by the GA UK grant No. 300823, the Charles University research program Cooperatio-Neurosciences. and a grant by the Programme Johannes Amos Comenius under the Ministry of Education, Youth and Sports of the Czech Republic, CZ.02.01.01/00/23_020/0008560 (“BRAINSCAPE: Cross-sectoral collaboration for the development of new technologies in psychiatry – precision medicine and new therapies”), and the Czech Health Research Council (project AZV 17-31852A).

## Supplementary material

**Table S1.**
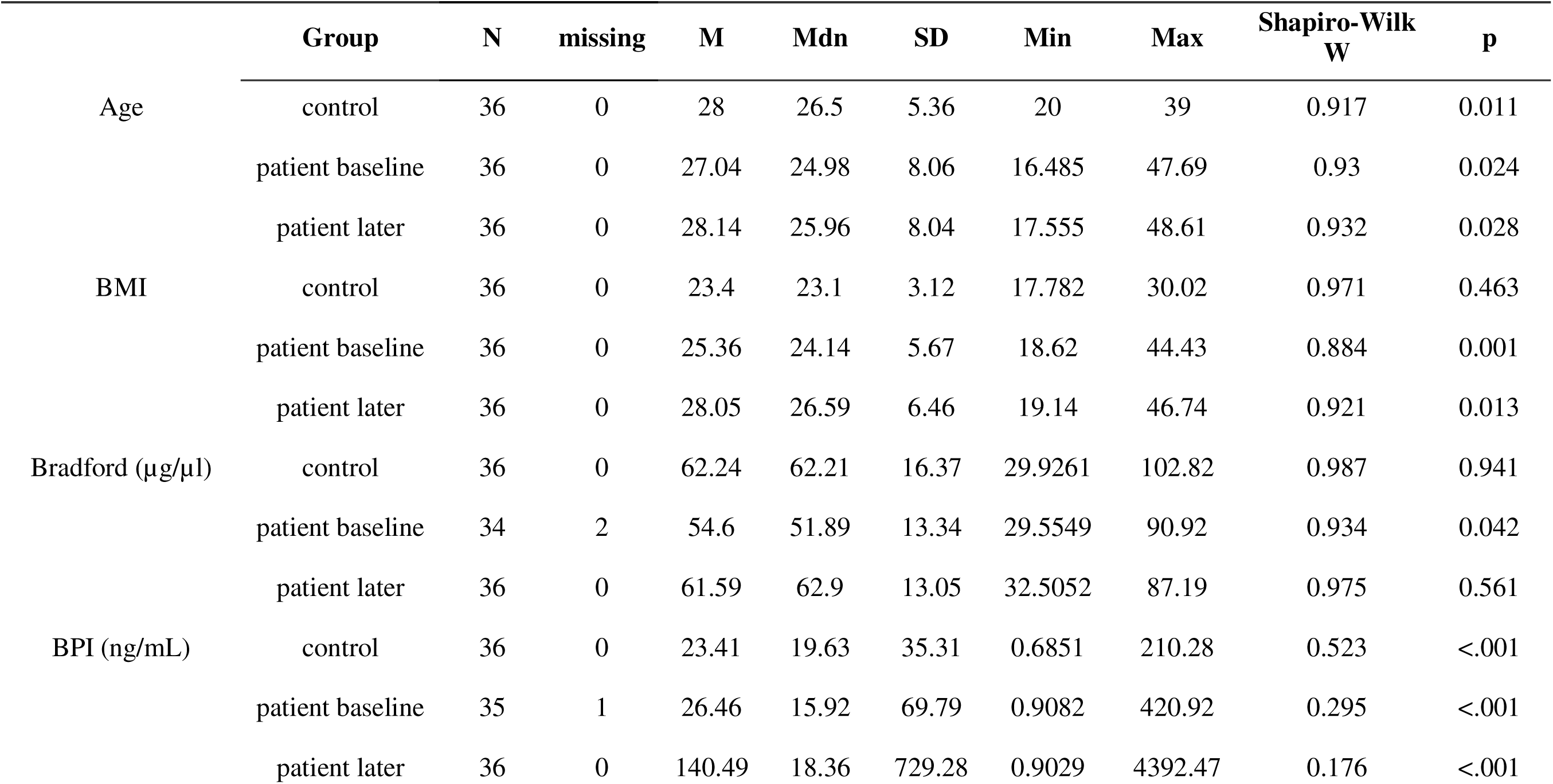
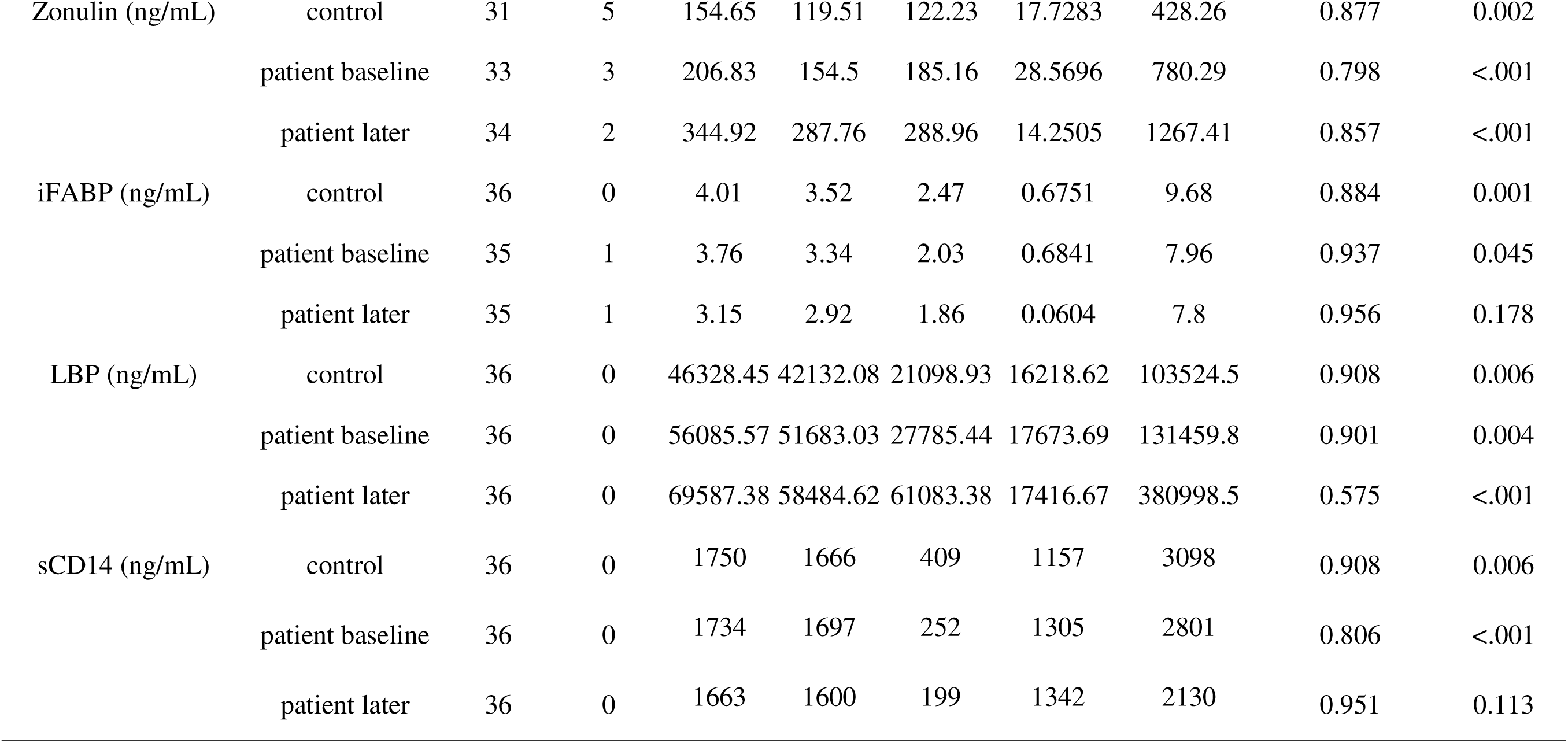
Descriptive analysis of the sample divided by groups.

## Sensitivity analyses

**Table S2a.**
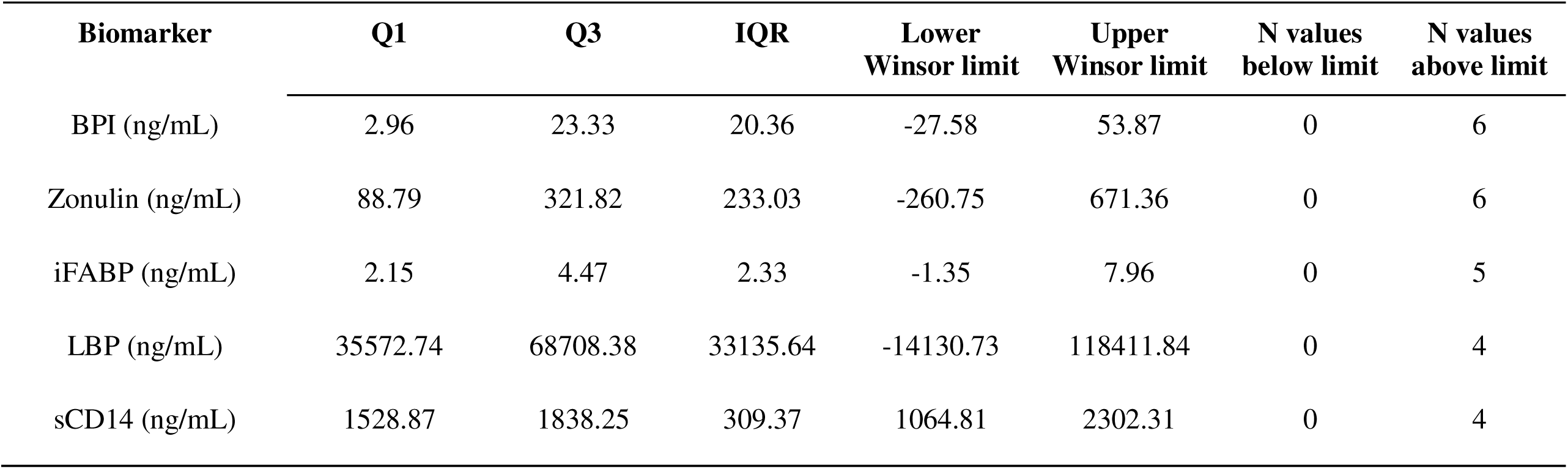
Winsorization thresholds.

**Table S2b.**
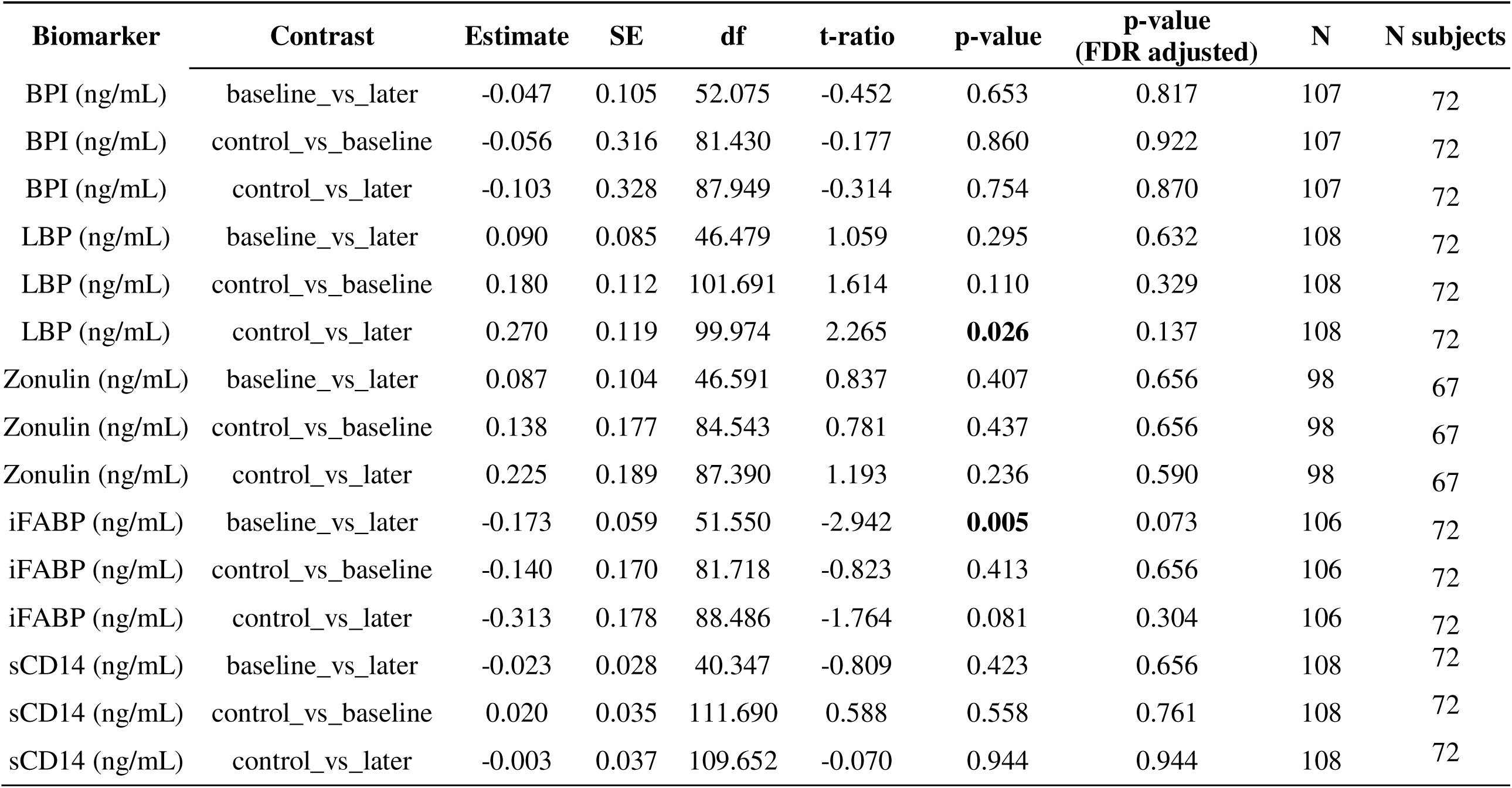
Summary data of comparisons, using Winsorized data.

**Table S3.**
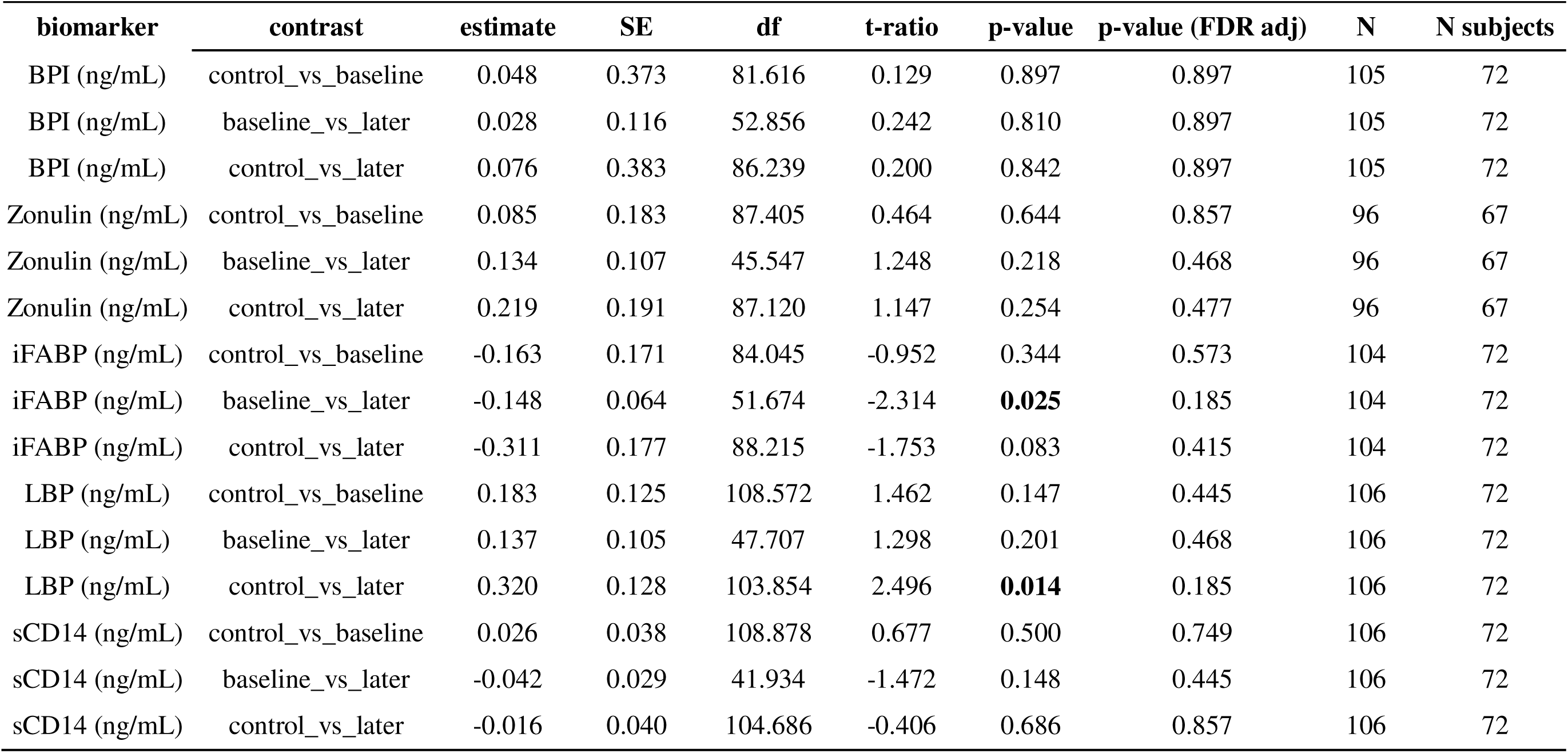
Bradford-adjusted LMM results.

**Table S4.**
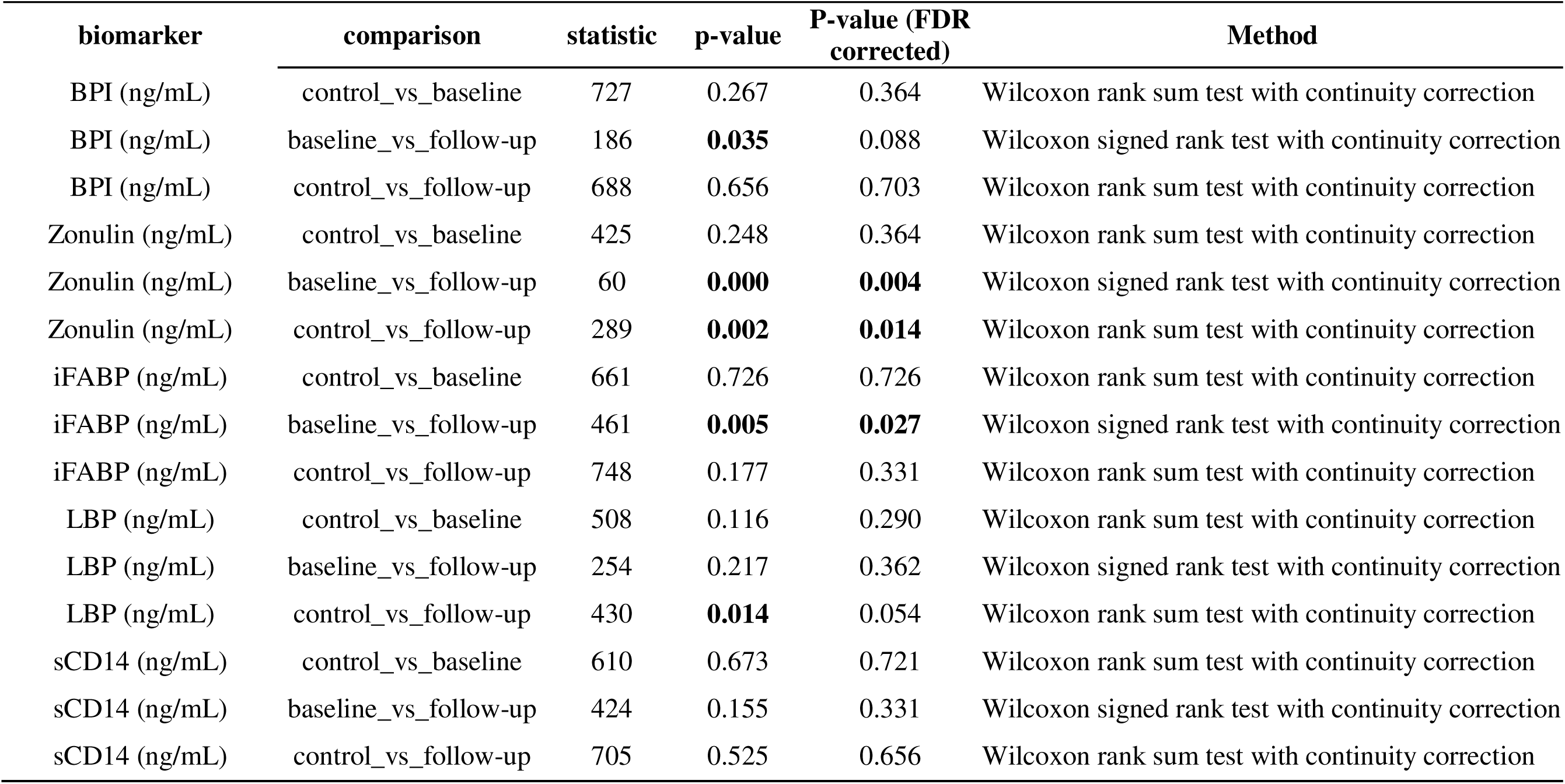
Wilcoxon test for sensitivity check.

**Table S5.**
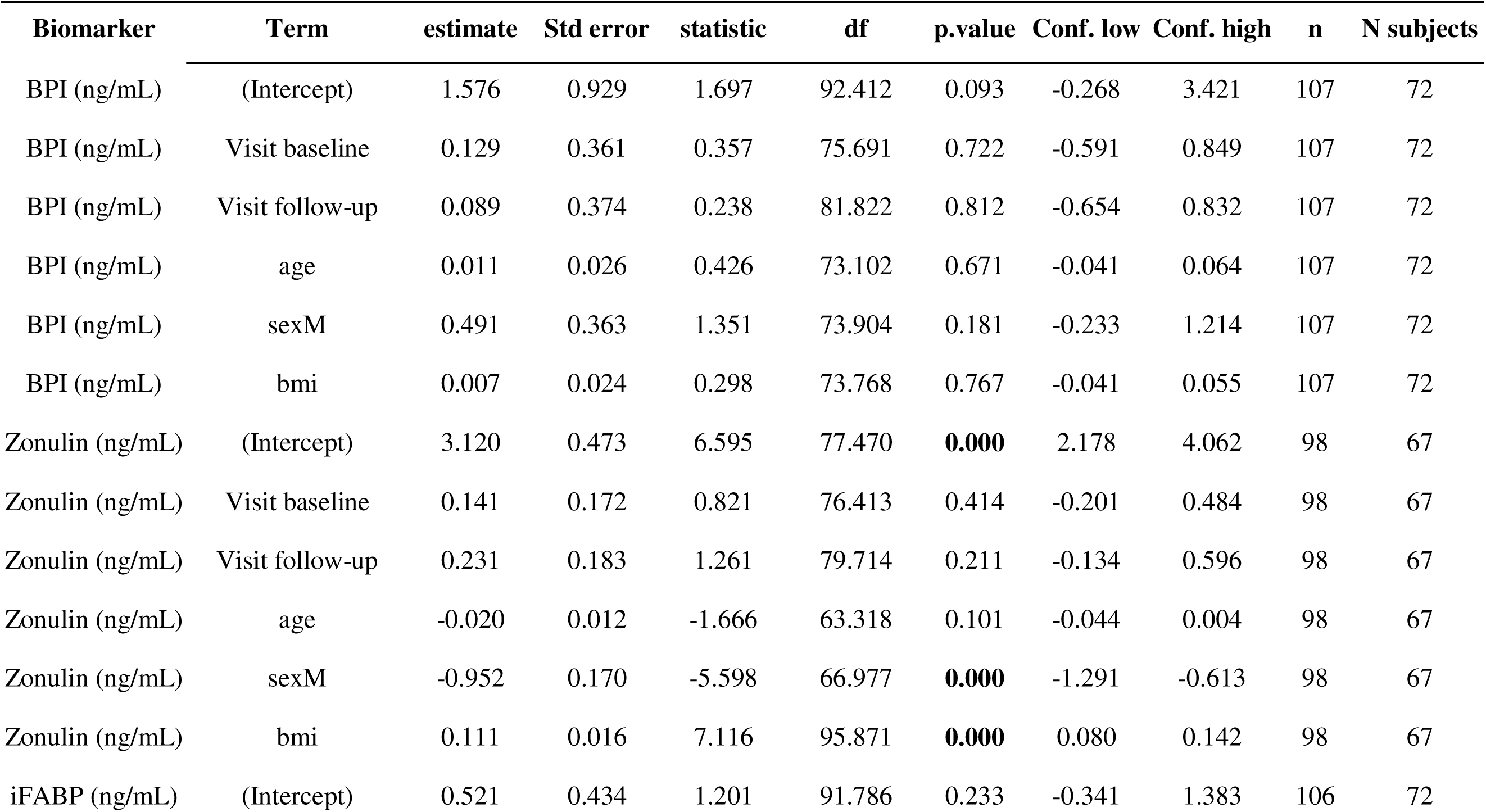
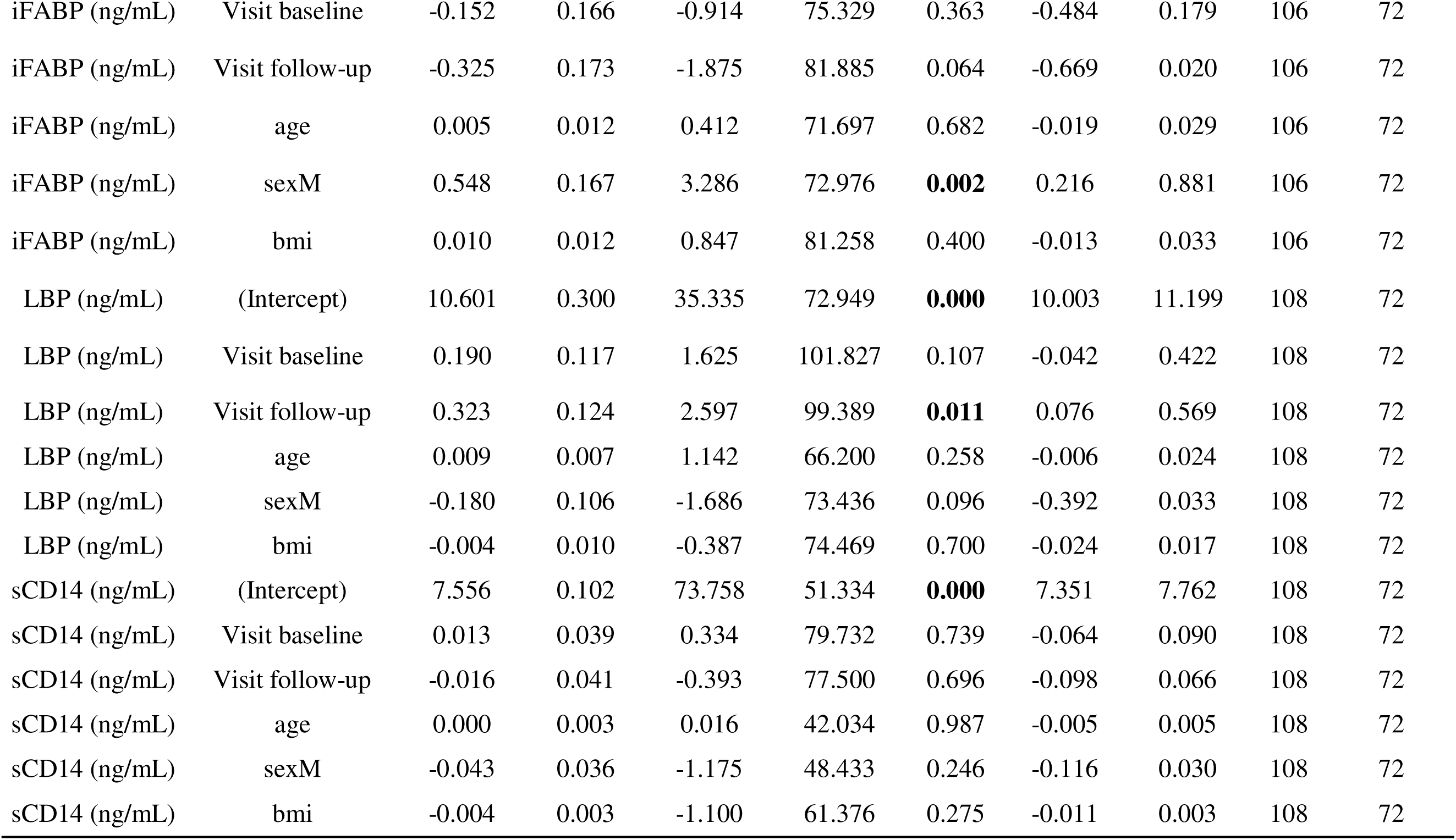
Primary fixed effects linear mixed model.

**Table S6.**
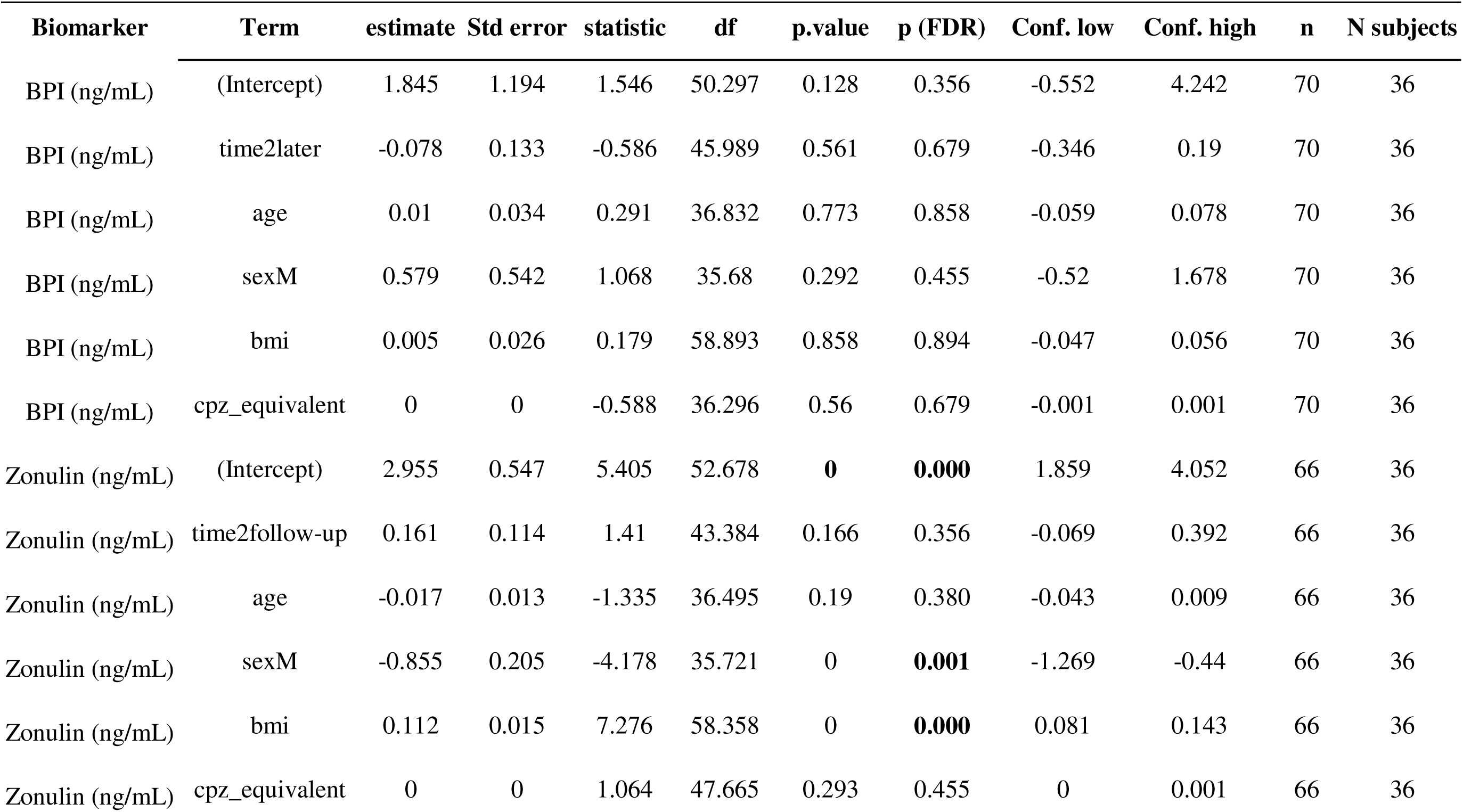
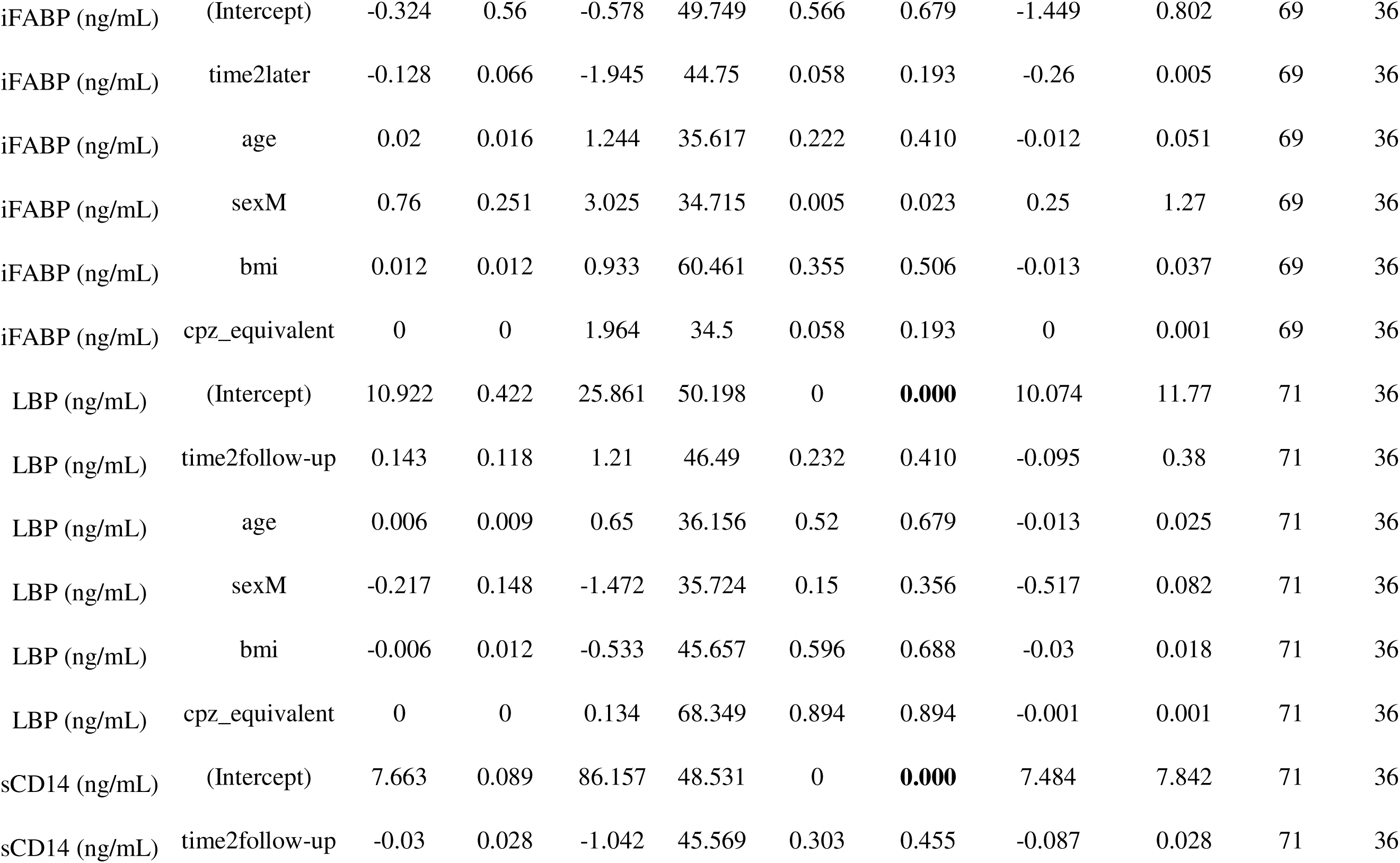
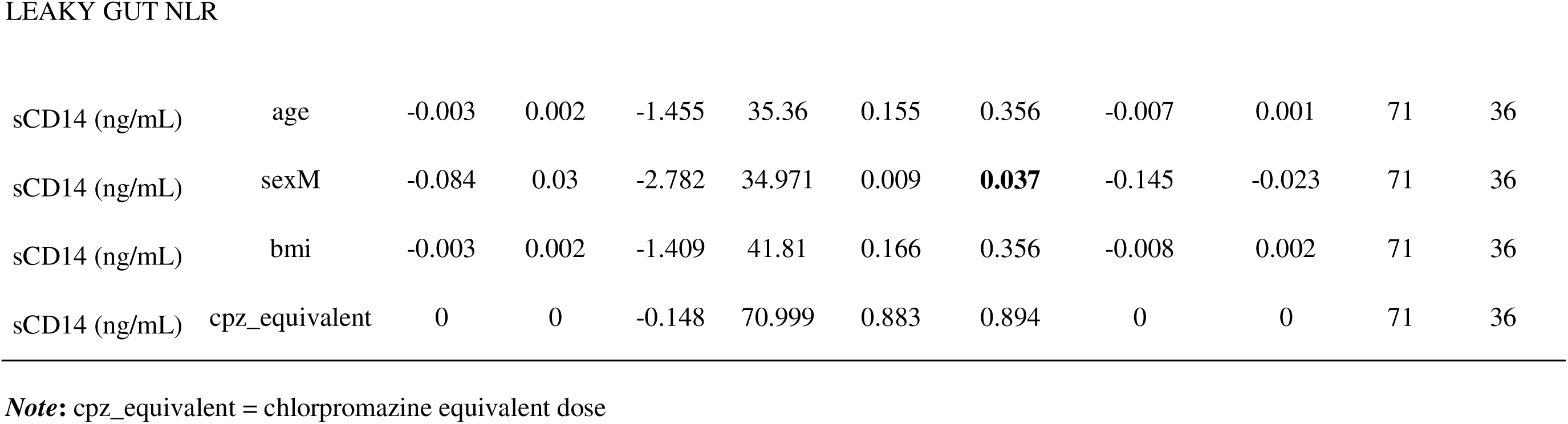
Primary fixed effects linear mixed model, adjusting for chlorpromazine exposure in the patient sample.

**Table S7.**
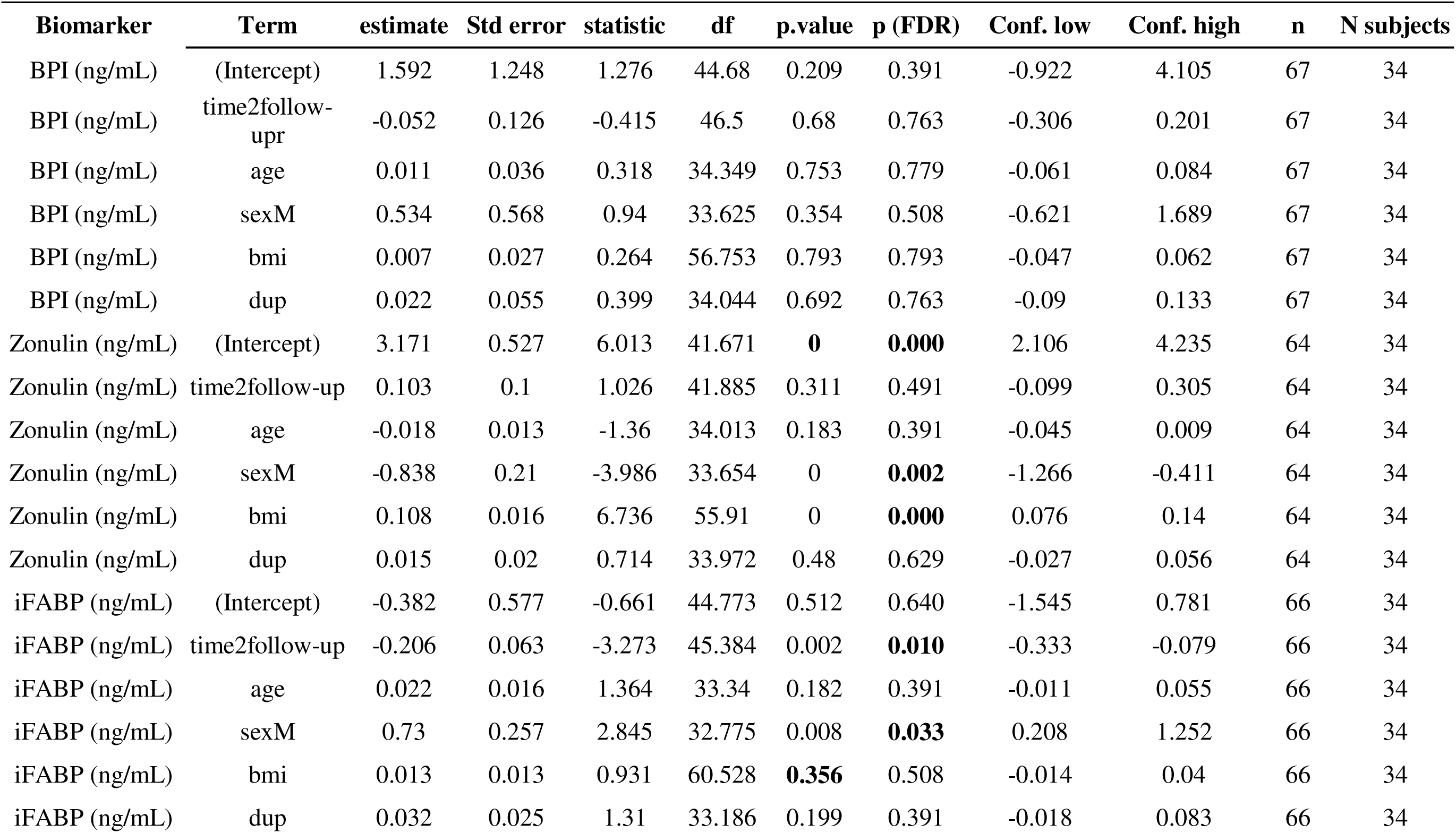
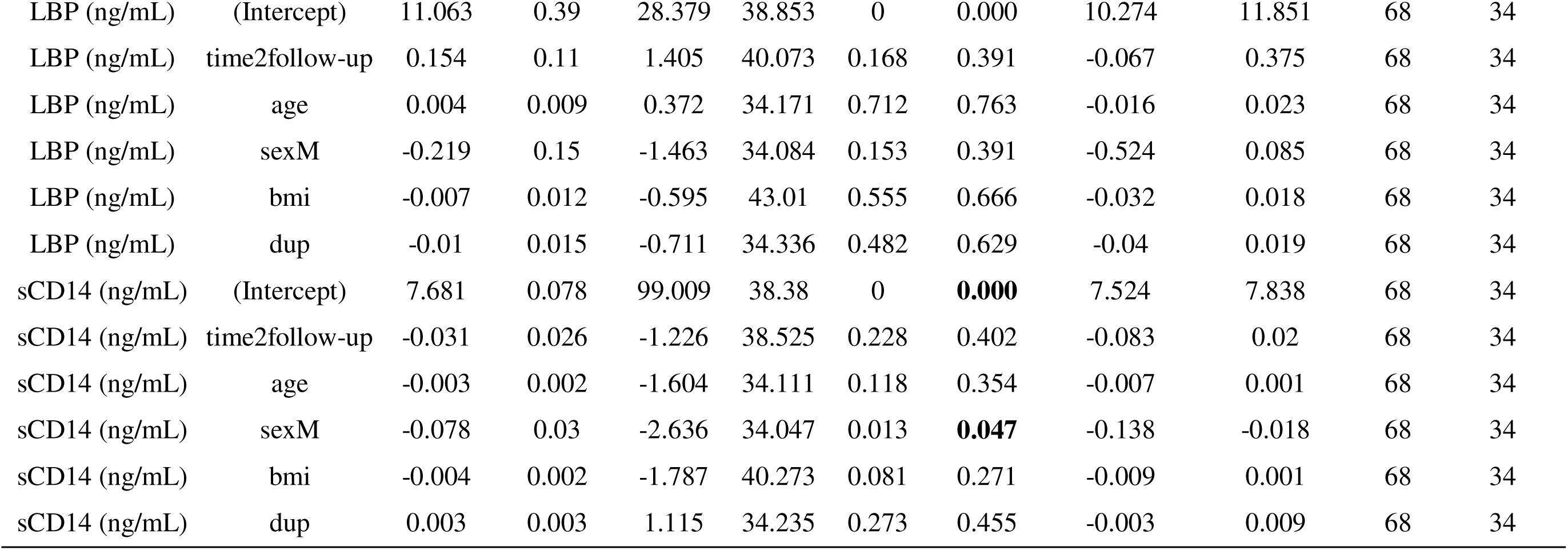
Primary fixed effects linear mixed model, adjusting for duration of untreated psychosis (DUP) in the patient sample.

**Supplementary Figure S1.**
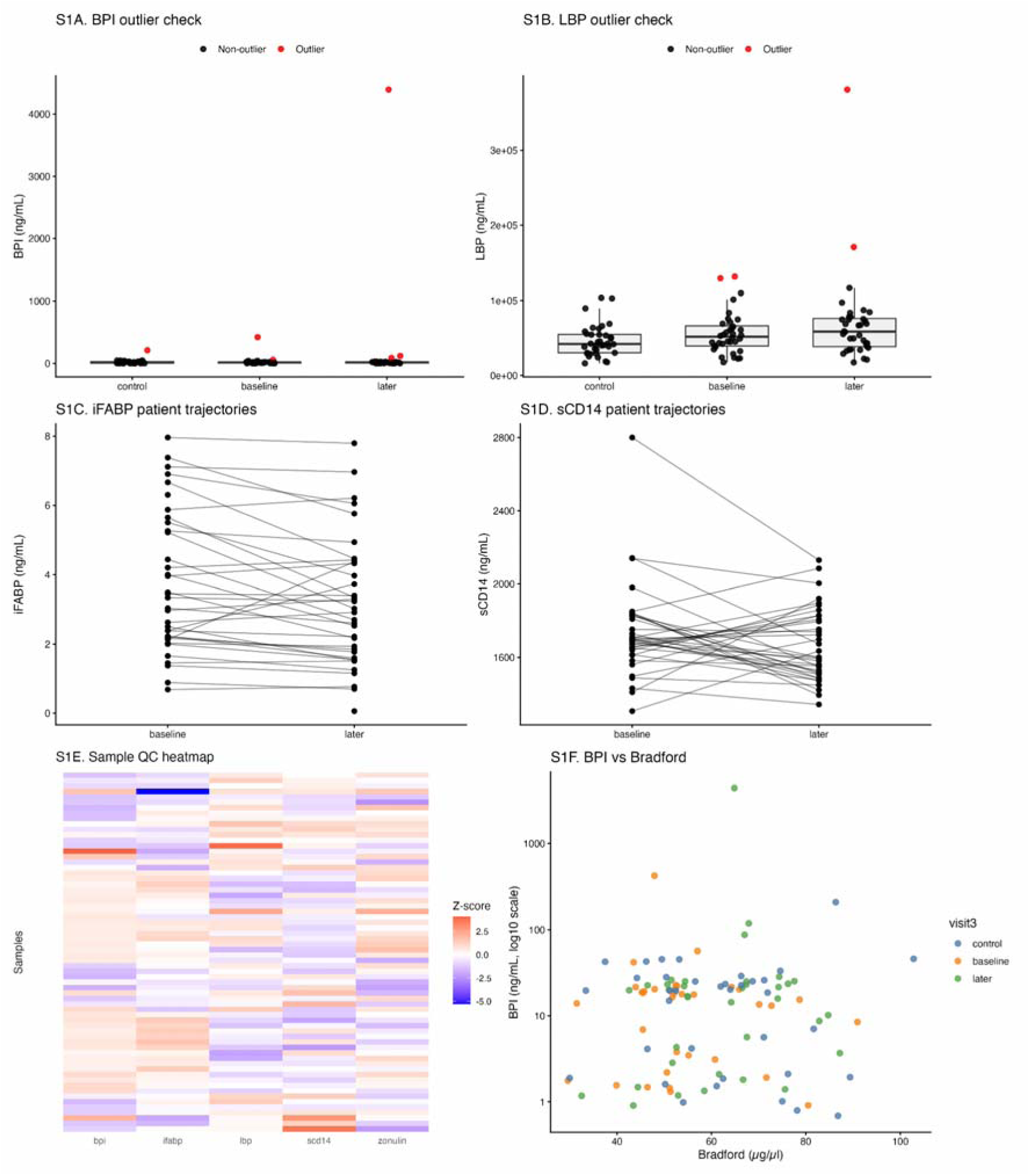
Quality control, outlier inspection, and sensitivity-related visualizations. **(A)** Outlier-highlighted distribution of BPI across controls, baseline patients, and follow-up patients. **(B)** Outlier-highlighted distribution of LBP across groups. **(C)** Within-patient longitudinal trajectories for iFABP. **(D)** Within-patient longitudinal trajectories for sCD14. **(E)** Sample-level heatmap of standardized log-transformed biomarker values across all complete cases. **(F)** Relationship between BPI and Bradford protein concentration, shown to assess whether extreme biomarker values were explained by total protein content

**Table S8.**
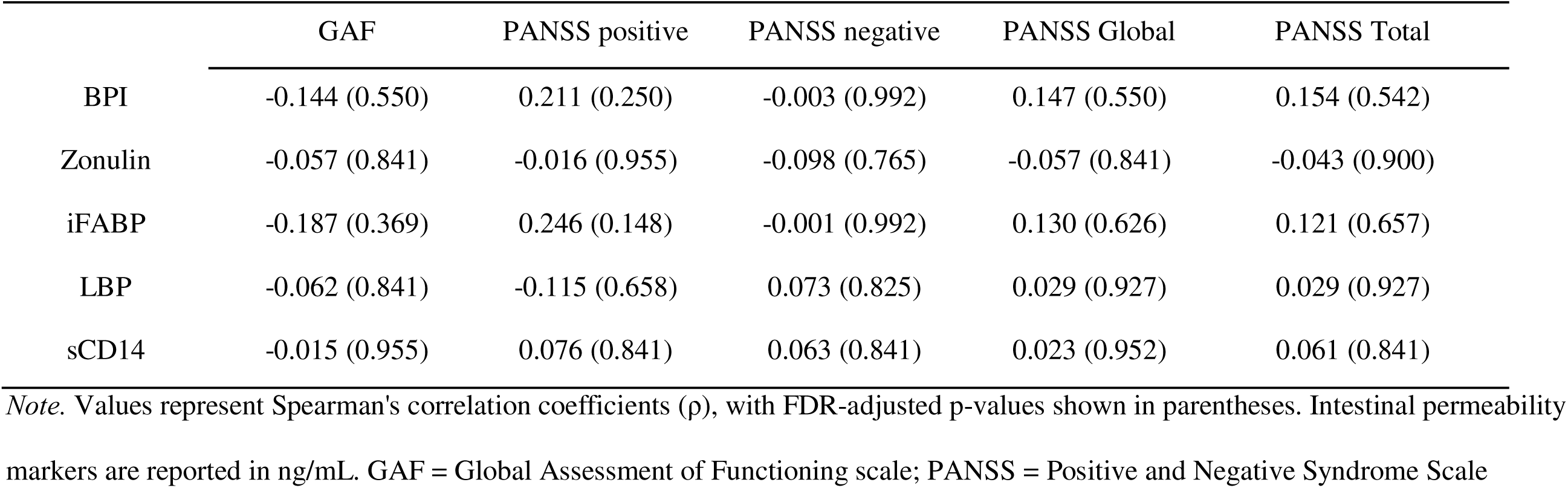
Spearman correlation coefficient (and FDR corrected p-value) between intestinal permeability indicators and clinical severity scales (only in the sample of patients, both time points)

**Table S9.**
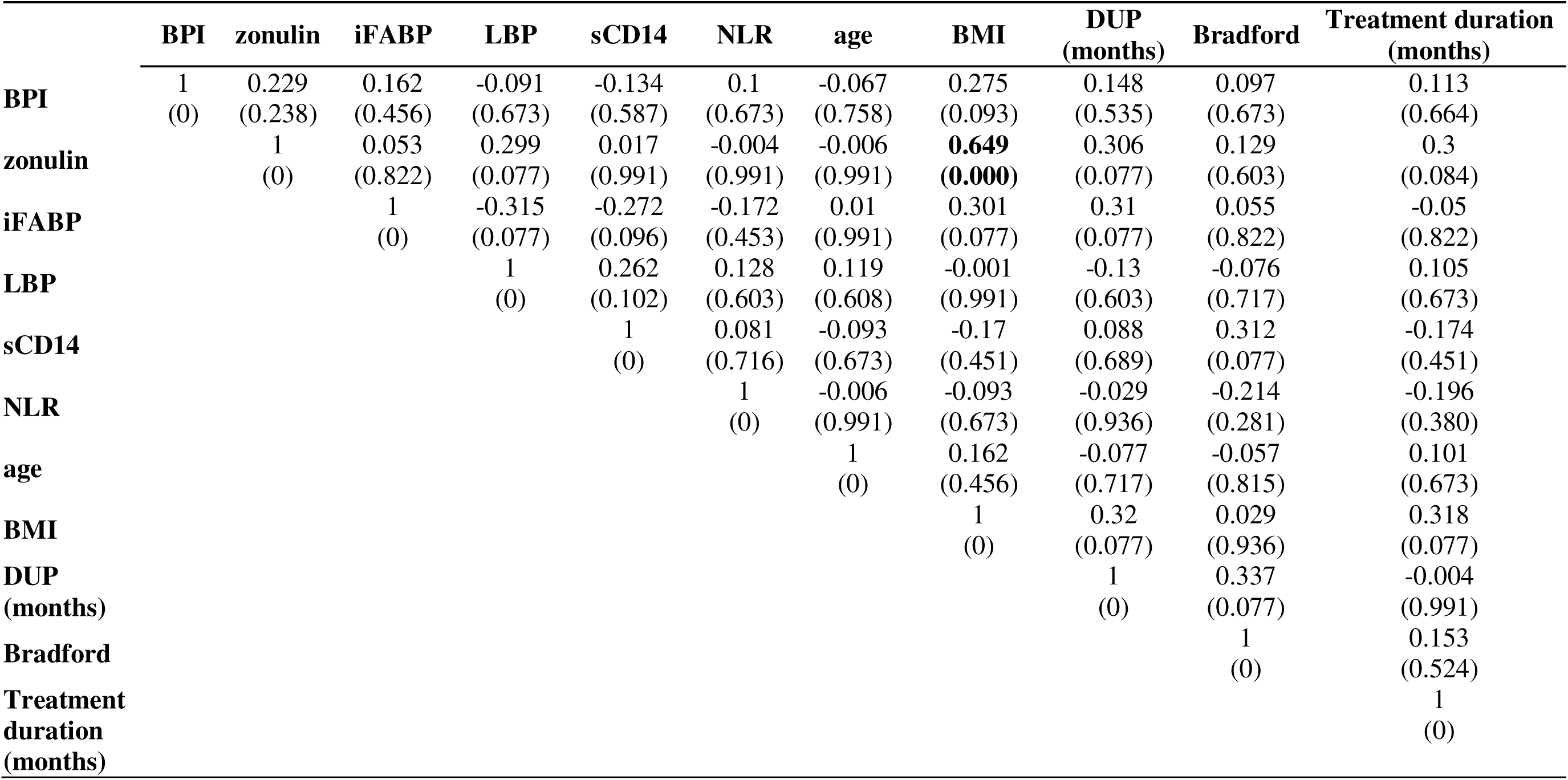
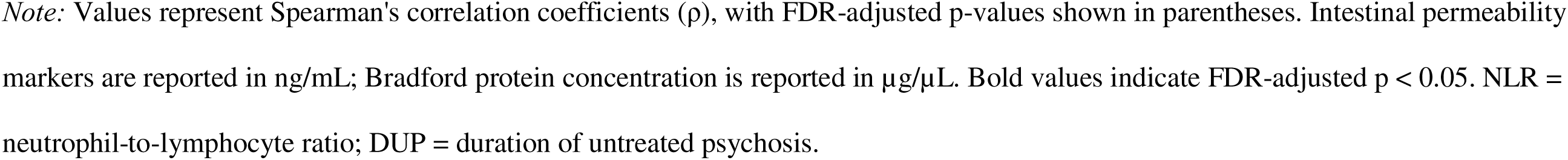
Spearman correlation coefficient (and FDR corrected p-value) between intestinal permeability markers and patient covariates (Age, BMI, duration of untreated psychosis, Bradford serum protein concentration)

