## Supplementary file for "From Gut to Brain: A Longitudinal Assessment of Intestinal Permeability and Immune Activation Markers in Early-Stage Schizophrenia Spectrum Disorders"

**Supplementary material**

**Table S1.** Descriptive analysis of the sample divided by groups

| | Group | N | missing | M | Mdn | SD | Min | Max | Shapiro-Wilk W | p |
|---|---|---|---|---|---|---|---|---|---|---|
| Age | control | 36 | 0 | 28 | 26.5 | 5.36 | 20 | 39 | 0.917 | 0.011 |
| | patient baseline | 36 | 0 | 27.04 | 24.98 | 8.06 | 16.485 | 47.69 | 0.93 | 0.024 |
| | patient later | 36 | 0 | 28.14 | 25.96 | 8.04 | 17.555 | 48.61 | 0.932 | 0.028 |
| BMI | control | 36 | 0 | 23.4 | 23.1 | 3.12 | 17.782 | 30.02 | 0.971 | 0.463 |
| | patient baseline | 36 | 0 | 25.36 | 24.14 | 5.67 | 18.62 | 44.43 | 0.884 | 0.001 |
| | patient later | 36 | 0 | 28.05 | 26.59 | 6.46 | 19.14 | 46.74 | 0.921 | 0.013 |
| Bradford (µg/µl) | control | 36 | 0 | 62.24 | 62.21 | 16.37 | 29.9261 | 102.82 | 0.987 | 0.941 |
| | patient baseline | 34 | 2 | 54.6 | 51.89 | 13.34 | 29.5549 | 90.92 | 0.934 | 0.042 |
| | patient later | 36 | 0 | 61.59 | 62.9 | 13.05 | 32.5052 | 87.19 | 0.975 | 0.561 |
| BPI (ng/mL) | control | 36 | 0 | 23.41 | 19.63 | 35.31 | 0.6851 | 210.28 | 0.523 | <.001 |
| | patient baseline | 35 | 1 | 26.46 | 15.92 | 69.79 | 0.9082 | 420.92 | 0.295 | <.001 |
| | patient later | 36 | 0 | 140.49 | 18.36 | 729.28 | 0.9029 | 4392.47 | 0.176 | <.001 |

| | | | | | | | | | | |
|---|---|---|---|---|---|---|---|---|---|---|
| Zonulin (ng/mL) | control | 31 | 5 | 154.65 | 119.51 | 122.23 | 17.7283 | 428.26 | 0.877 | 0.002 |
| | patient baseline | 33 | 3 | 206.83 | 154.5 | 185.16 | 28.5696 | 780.29 | 0.798 | <.001 |
| | patient later | 34 | 2 | 344.92 | 287.76 | 288.96 | 14.2505 | 1267.41 | 0.857 | <.001 |
| iFABP (ng/mL) | control | 36 | 0 | 4.01 | 3.52 | 2.47 | 0.6751 | 9.68 | 0.884 | 0.001 |
| | patient baseline | 35 | 1 | 3.76 | 3.34 | 2.03 | 0.6841 | 7.96 | 0.937 | 0.045 |
| | patient later | 35 | 1 | 3.15 | 2.92 | 1.86 | 0.0604 | 7.8 | 0.956 | 0.178 |
| LBP (ng/mL) | control | 36 | 0 | 46328.45 | 42132.08 | 21098.93 | 16218.62 | 103524.5 | 0.908 | 0.006 |
| | patient baseline | 36 | 0 | 56085.57 | 51683.03 | 27785.44 | 17673.69 | 131459.8 | 0.901 | 0.004 |
| | patient later | 36 | 0 | 69587.38 | 58484.62 | 61083.38 | 17416.67 | 380998.5 | 0.575 | <.001 |
| sCD14 (ng/mL) | control | 36 | 0 | 1750 | 1666 | 409 | 1157 | 3098 | 0.908 | 0.006 |
| | patient baseline | 36 | 0 | 1734 | 1697 | 252 | 1305 | 2801 | 0.806 | <.001 |
| | patient later | 36 | 0 | 1663 | 1600 | 199 | 1342 | 2130 | 0.951 | 0.113 |

**Sensitivity analyses**

**Table S2a.** Winsorization thresholds

| Biomarker | Q1 | Q3 | IQR | Lower Winsor limit | Upper Winsor limit | N values below limit | N values above limit |
|---|---|---|---|---|---|---|---|
| BPI (ng/mL) | 2.96 | 23.33 | 20.36 | -27.58 | 53.87 | 0 | 6 |
| Zonulin (ng/mL) | 88.79 | 321.82 | 233.03 | -260.75 | 671.36 | 0 | 6 |
| iFABP (ng/mL) | 2.15 | 4.47 | 2.33 | -1.35 | 7.96 | 0 | 5 |
| LBP (ng/mL) | 35572.74 | 68708.38 | 33135.64 | -14130.73 | 118411.84 | 0 | 4 |
| sCD14 (ng/mL) | 1528.87 | 1838.25 | 309.37 | 1064.81 | 2302.31 | 0 | 4 |

**Table S2b.** Summary data of comparisons, using Winsorized data

| Biomarker | Contrast | Estimate | SE | df | t-ratio | p-value | p-value (FDR adjusted) | N | N subjects |
|---|---|---|---|---|---|---|---|---|---|
| BPI (ng/mL) | baseline_vs_later | -0.047 | 0.105 | 52.075 | -0.452 | 0.653 | 0.817 | 107 | 72 |
| BPI (ng/mL) | control_vs_baseline | -0.056 | 0.316 | 81.430 | -0.177 | 0.860 | 0.922 | 107 | 72 |
| BPI (ng/mL) | control_vs_later | -0.103 | 0.328 | 87.949 | -0.314 | 0.754 | 0.870 | 107 | 72 |
| LBP (ng/mL) | baseline_vs_later | 0.090 | 0.085 | 46.479 | 1.059 | 0.295 | 0.632 | 108 | 72 |
| LBP (ng/mL) | control_vs_baseline | 0.180 | 0.112 | 101.691 | 1.614 | 0.110 | 0.329 | 108 | 72 |
| LBP (ng/mL) | control_vs_later | 0.270 | 0.119 | 99.974 | 2.265 | **0.026** | 0.137 | 108 | 72 |
| Zonulin (ng/mL) | baseline_vs_later | 0.087 | 0.104 | 46.591 | 0.837 | 0.407 | 0.656 | 98 | 67 |
| Zonulin (ng/mL) | control_vs_baseline | 0.138 | 0.177 | 84.543 | 0.781 | 0.437 | 0.656 | 98 | 67 |
| Zonulin (ng/mL) | control_vs_later | 0.225 | 0.189 | 87.390 | 1.193 | 0.236 | 0.590 | 98 | 67 |
| iFABP (ng/mL) | baseline_vs_later | -0.173 | 0.059 | 51.550 | -2.942 | **0.005** | 0.073 | 106 | 72 |
| iFABP (ng/mL) | control_vs_baseline | -0.140 | 0.170 | 81.718 | -0.823 | 0.413 | 0.656 | 106 | 72 |
| iFABP (ng/mL) | control_vs_later | -0.313 | 0.178 | 88.486 | -1.764 | 0.081 | 0.304 | 106 | 72 |
| sCD14 (ng/mL) | baseline_vs_later | -0.023 | 0.028 | 40.347 | -0.809 | 0.423 | 0.656 | 108 | 72 |
| sCD14 (ng/mL) | control_vs_baseline | 0.020 | 0.035 | 111.690 | 0.588 | 0.558 | 0.761 | 108 | 72 |
| sCD14 (ng/mL) | control_vs_later | -0.003 | 0.037 | 109.652 | -0.070 | 0.944 | 0.944 | 108 | 72 |

**Table S3.** Bradford-adjusted LMM results

| biomarker | contrast | estimate | SE | df | t-ratio | p-value | p-value (FDR adj) | N | N subjects |
|---|---|---|---|---|---|---|---|---|---|
| BPI (ng/mL) | control_vs_baseline | 0.048 | 0.373 | 81.616 | 0.129 | 0.897 | 0.897 | 105 | 72 |
| BPI (ng/mL) | baseline_vs_later | 0.028 | 0.116 | 52.856 | 0.242 | 0.810 | 0.897 | 105 | 72 |
| BPI (ng/mL) | control_vs_later | 0.076 | 0.383 | 86.239 | 0.200 | 0.842 | 0.897 | 105 | 72 |
| Zonulin (ng/mL) | control_vs_baseline | 0.085 | 0.183 | 87.405 | 0.464 | 0.644 | 0.857 | 96 | 67 |
| Zonulin (ng/mL) | baseline_vs_later | 0.134 | 0.107 | 45.547 | 1.248 | 0.218 | 0.468 | 96 | 67 |
| Zonulin (ng/mL) | control_vs_later | 0.219 | 0.191 | 87.120 | 1.147 | 0.254 | 0.477 | 96 | 67 |
| iFABP (ng/mL) | control_vs_baseline | -0.163 | 0.171 | 84.045 | -0.952 | 0.344 | 0.573 | 104 | 72 |
| iFABP (ng/mL) | baseline_vs_later | -0.148 | 0.064 | 51.674 | -2.314 | **0.025** | 0.185 | 104 | 72 |
| iFABP (ng/mL) | control_vs_later | -0.311 | 0.177 | 88.215 | -1.753 | 0.083 | 0.415 | 104 | 72 |
| LBP (ng/mL) | control_vs_baseline | 0.183 | 0.125 | 108.572 | 1.462 | 0.147 | 0.445 | 106 | 72 |
| LBP (ng/mL) | baseline_vs_later | 0.137 | 0.105 | 47.707 | 1.298 | 0.201 | 0.468 | 106 | 72 |
| LBP (ng/mL) | control_vs_later | 0.320 | 0.128 | 103.854 | 2.496 | **0.014** | 0.185 | 106 | 72 |
| sCD14 (ng/mL) | control_vs_baseline | 0.026 | 0.038 | 108.878 | 0.677 | 0.500 | 0.749 | 106 | 72 |
| sCD14 (ng/mL) | baseline_vs_later | -0.042 | 0.029 | 41.934 | -1.472 | 0.148 | 0.445 | 106 | 72 |
| sCD14 (ng/mL) | control_vs_later | -0.016 | 0.040 | 104.686 | -0.406 | 0.686 | 0.857 | 106 | 72 |

**Table S4.** Wilcoxon test for sensitivity check

| biomarker | comparison | statistic | p-value | P-value (FDR corrected) | Method |
|---|---|---|---|---|---|
| BPI (ng/mL) | control_vs_baseline | 727 | 0.267 | 0.364 | Wilcoxon rank sum test with continuity correction |
| BPI (ng/mL) | baseline_vs_follow-up | 186 | **0.035** | 0.088 | Wilcoxon signed rank test with continuity correction |
| BPI (ng/mL) | control_vs_follow-up | 688 | 0.656 | 0.703 | Wilcoxon rank sum test with continuity correction |
| Zonulin (ng/mL) | control_vs_baseline | 425 | 0.248 | 0.364 | Wilcoxon rank sum test with continuity correction |
| Zonulin (ng/mL) | baseline_vs_follow-up | 60 | **0.000** | **0.004** | Wilcoxon signed rank test with continuity correction |
| Zonulin (ng/mL) | control_vs_follow-up | 289 | **0.002** | **0.014** | Wilcoxon rank sum test with continuity correction |
| iFABP (ng/mL) | control_vs_baseline | 661 | 0.726 | 0.726 | Wilcoxon rank sum test with continuity correction |
| iFABP (ng/mL) | baseline_vs_follow-up | 461 | **0.005** | **0.027** | Wilcoxon signed rank test with continuity correction |
| iFABP (ng/mL) | control_vs_follow-up | 748 | 0.177 | 0.331 | Wilcoxon rank sum test with continuity correction |
| LBP (ng/mL) | control_vs_baseline | 508 | 0.116 | 0.290 | Wilcoxon rank sum test with continuity correction |
| LBP (ng/mL) | baseline_vs_follow-up | 254 | 0.217 | 0.362 | Wilcoxon signed rank test with continuity correction |
| LBP (ng/mL) | control_vs_follow-up | 430 | **0.014** | 0.054 | Wilcoxon rank sum test with continuity correction |
| sCD14 (ng/mL) | control_vs_baseline | 610 | 0.673 | 0.721 | Wilcoxon rank sum test with continuity correction |
| sCD14 (ng/mL) | baseline_vs_follow-up | 424 | 0.155 | 0.331 | Wilcoxon signed rank test with continuity correction |
| sCD14 (ng/mL) | control_vs_follow-up | 705 | 0.525 | 0.656 | Wilcoxon rank sum test with continuity correction |

**Table S5.** Primary fixed effects linear mixed model

| Biomarker | Term | estimate | Std error | statistic | df | p.value | Conf. low | Conf. high | n | N subjects |
|---|---|---|---|---|---|---|---|---|---|---|
| BPI (ng/mL) | (Intercept) | 1.576 | 0.929 | 1.697 | 92.412 | 0.093 | -0.268 | 3.421 | 107 | 72 |
| BPI (ng/mL) | Visit baseline | 0.129 | 0.361 | 0.357 | 75.691 | 0.722 | -0.591 | 0.849 | 107 | 72 |
| BPI (ng/mL) | Visit follow-up | 0.089 | 0.374 | 0.238 | 81.822 | 0.812 | -0.654 | 0.832 | 107 | 72 |
| BPI (ng/mL) | age | 0.011 | 0.026 | 0.426 | 73.102 | 0.671 | -0.041 | 0.064 | 107 | 72 |
| BPI (ng/mL) | sexM | 0.491 | 0.363 | 1.351 | 73.904 | 0.181 | -0.233 | 1.214 | 107 | 72 |
| BPI (ng/mL) | bmi | 0.007 | 0.024 | 0.298 | 73.768 | 0.767 | -0.041 | 0.055 | 107 | 72 |
| Zonulin (ng/mL) | (Intercept) | 3.120 | 0.473 | 6.595 | 77.470 | **0.000** | 2.178 | 4.062 | 98 | 67 |
| Zonulin (ng/mL) | Visit baseline | 0.141 | 0.172 | 0.821 | 76.413 | 0.414 | -0.201 | 0.484 | 98 | 67 |
| Zonulin (ng/mL) | Visit follow-up | 0.231 | 0.183 | 1.261 | 79.714 | 0.211 | -0.134 | 0.596 | 98 | 67 |
| Zonulin (ng/mL) | age | -0.020 | 0.012 | -1.666 | 63.318 | 0.101 | -0.044 | 0.004 | 98 | 67 |
| Zonulin (ng/mL) | sexM | -0.952 | 0.170 | -5.598 | 66.977 | **0.000** | -1.291 | -0.613 | 98 | 67 |
| Zonulin (ng/mL) | bmi | 0.111 | 0.016 | 7.116 | 95.871 | **0.000** | 0.080 | 0.142 | 98 | 67 |
| iFABP (ng/mL) | (Intercept) | 0.521 | 0.434 | 1.201 | 91.786 | 0.233 | -0.341 | 1.383 | 106 | 72 |

| | | | | | | | | | | |
|---|---|---|---|---|---|---|---|---|---|---|
| iFABP (ng/mL) | Visit baseline | -0.152 | 0.166 | -0.914 | 75.329 | 0.363 | -0.484 | 0.179 | 106 | 72 |
| iFABP (ng/mL) | Visit follow-up | -0.325 | 0.173 | -1.875 | 81.885 | 0.064 | -0.669 | 0.020 | 106 | 72 |
| iFABP (ng/mL) | age | 0.005 | 0.012 | 0.412 | 71.697 | 0.682 | -0.019 | 0.029 | 106 | 72 |
| iFABP (ng/mL) | sexM | 0.548 | 0.167 | 3.286 | 72.976 | **0.002** | 0.216 | 0.881 | 106 | 72 |
| iFABP (ng/mL) | bmi | 0.010 | 0.012 | 0.847 | 81.258 | 0.400 | -0.013 | 0.033 | 106 | 72 |
| LBP (ng/mL) | (Intercept) | 10.601 | 0.300 | 35.335 | 72.949 | **0.000** | 10.003 | 11.199 | 108 | 72 |
| LBP (ng/mL) | Visit baseline | 0.190 | 0.117 | 1.625 | 101.827 | 0.107 | -0.042 | 0.422 | 108 | 72 |
| LBP (ng/mL) | Visit follow-up | 0.323 | 0.124 | 2.597 | 99.389 | **0.011** | 0.076 | 0.569 | 108 | 72 |
| LBP (ng/mL) | age | 0.009 | 0.007 | 1.142 | 66.200 | 0.258 | -0.006 | 0.024 | 108 | 72 |
| LBP (ng/mL) | sexM | -0.180 | 0.106 | -1.686 | 73.436 | 0.096 | -0.392 | 0.033 | 108 | 72 |
| LBP (ng/mL) | bmi | -0.004 | 0.010 | -0.387 | 74.469 | 0.700 | -0.024 | 0.017 | 108 | 72 |
| sCD14 (ng/mL) | (Intercept) | 7.556 | 0.102 | 73.758 | 51.334 | **0.000** | 7.351 | 7.762 | 108 | 72 |
| sCD14 (ng/mL) | Visit baseline | 0.013 | 0.039 | 0.334 | 79.732 | 0.739 | -0.064 | 0.090 | 108 | 72 |
| sCD14 (ng/mL) | Visit follow-up | -0.016 | 0.041 | -0.393 | 77.500 | 0.696 | -0.098 | 0.066 | 108 | 72 |
| sCD14 (ng/mL) | age | 0.000 | 0.003 | 0.016 | 42.034 | 0.987 | -0.005 | 0.005 | 108 | 72 |
| sCD14 (ng/mL) | sexM | -0.043 | 0.036 | -1.175 | 48.433 | 0.246 | -0.116 | 0.030 | 108 | 72 |
| sCD14 (ng/mL) | bmi | -0.004 | 0.003 | -1.100 | 61.376 | 0.275 | -0.011 | 0.003 | 108 | 72 |

**Table S6.** Primary fixed effects linear mixed model, adjusting for chlorpromazine exposure in the patient sample

| Biomarker | Term | estimate | Std error | statistic | df | p.value | p (FDR) | Conf. low | Conf. high | n | N subjects |
|---|---|---|---|---|---|---|---|---|---|---|---|
| BPI (ng/mL) | (Intercept) | 1.845 | 1.194 | 1.546 | 50.297 | 0.128 | 0.356 | -0.552 | 4.242 | 70 | 36 |
| BPI (ng/mL) | time2later | -0.078 | 0.133 | -0.586 | 45.989 | 0.561 | 0.679 | -0.346 | 0.19 | 70 | 36 |
| BPI (ng/mL) | age | 0.01 | 0.034 | 0.291 | 36.832 | 0.773 | 0.858 | -0.059 | 0.078 | 70 | 36 |
| BPI (ng/mL) | sexM | 0.579 | 0.542 | 1.068 | 35.68 | 0.292 | 0.455 | -0.52 | 1.678 | 70 | 36 |
| BPI (ng/mL) | bmi | 0.005 | 0.026 | 0.179 | 58.893 | 0.858 | 0.894 | -0.047 | 0.056 | 70 | 36 |
| BPI (ng/mL) | cpz_equivalent | 0 | 0 | -0.588 | 36.296 | 0.56 | 0.679 | -0.001 | 0.001 | 70 | 36 |
| Zonulin (ng/mL) | (Intercept) | 2.955 | 0.547 | 5.405 | 52.678 | **0** | **0.000** | 1.859 | 4.052 | 66 | 36 |
| Zonulin (ng/mL) | time2follow-up | 0.161 | 0.114 | 1.41 | 43.384 | 0.166 | 0.356 | -0.069 | 0.392 | 66 | 36 |
| Zonulin (ng/mL) | age | -0.017 | 0.013 | -1.335 | 36.495 | 0.19 | 0.380 | -0.043 | 0.009 | 66 | 36 |
| Zonulin (ng/mL) | sexM | -0.855 | 0.205 | -4.178 | 35.721 | 0 | **0.001** | -1.269 | -0.44 | 66 | 36 |
| Zonulin (ng/mL) | bmi | 0.112 | 0.015 | 7.276 | 58.358 | 0 | **0.000** | 0.081 | 0.143 | 66 | 36 |
| Zonulin (ng/mL) | cpz_equivalent | 0 | 0 | 1.064 | 47.665 | 0.293 | 0.455 | 0 | 0.001 | 66 | 36 |

| | | | | | | | | | | | |
|---|---|---|---|---|---|---|---|---|---|---|---|
| iFABP (ng/mL) | (Intercept) | -0.324 | 0.56 | -0.578 | 49.749 | 0.566 | 0.679 | -1.449 | 0.802 | 69 | 36 |
| iFABP (ng/mL) | time2later | -0.128 | 0.066 | -1.945 | 44.75 | 0.058 | 0.193 | -0.26 | 0.005 | 69 | 36 |
| iFABP (ng/mL) | age | 0.02 | 0.016 | 1.244 | 35.617 | 0.222 | 0.410 | -0.012 | 0.051 | 69 | 36 |
| iFABP (ng/mL) | sexM | 0.76 | 0.251 | 3.025 | 34.715 | 0.005 | 0.023 | 0.25 | 1.27 | 69 | 36 |
| iFABP (ng/mL) | bmi | 0.012 | 0.012 | 0.933 | 60.461 | 0.355 | 0.506 | -0.013 | 0.037 | 69 | 36 |
| iFABP (ng/mL) | cpz_equivalent | 0 | 0 | 1.964 | 34.5 | 0.058 | 0.193 | 0 | 0.001 | 69 | 36 |
| LBP (ng/mL) | (Intercept) | 10.922 | 0.422 | 25.861 | 50.198 | 0 | **0.000** | 10.074 | 11.77 | 71 | 36 |
| LBP (ng/mL) | time2follow-up | 0.143 | 0.118 | 1.21 | 46.49 | 0.232 | 0.410 | -0.095 | 0.38 | 71 | 36 |
| LBP (ng/mL) | age | 0.006 | 0.009 | 0.65 | 36.156 | 0.52 | 0.679 | -0.013 | 0.025 | 71 | 36 |
| LBP (ng/mL) | sexM | -0.217 | 0.148 | -1.472 | 35.724 | 0.15 | 0.356 | -0.517 | 0.082 | 71 | 36 |
| LBP (ng/mL) | bmi | -0.006 | 0.012 | -0.533 | 45.657 | 0.596 | 0.688 | -0.03 | 0.018 | 71 | 36 |
| LBP (ng/mL) | cpz_equivalent | 0 | 0 | 0.134 | 68.349 | 0.894 | 0.894 | -0.001 | 0.001 | 71 | 36 |
| sCD14 (ng/mL) | (Intercept) | 7.663 | 0.089 | 86.157 | 48.531 | 0 | **0.000** | 7.484 | 7.842 | 71 | 36 |
| sCD14 (ng/mL) | time2follow-up | -0.03 | 0.028 | -1.042 | 45.569 | 0.303 | 0.455 | -0.087 | 0.028 | 71 | 36 |

| | | | | | | | | | | | |
|---|---|---|---|---|---|---|---|---|---|---|---|
| sCD14 (ng/mL) | age | -0.003 | 0.002 | -1.455 | 35.36 | 0.155 | 0.356 | -0.007 | 0.001 | 71 | 36 |
| sCD14 (ng/mL) | sexM | -0.084 | 0.03 | -2.782 | 34.971 | 0.009 | **0.037** | -0.145 | -0.023 | 71 | 36 |
| sCD14 (ng/mL) | bmi | -0.003 | 0.002 | -1.409 | 41.81 | 0.166 | 0.356 | -0.008 | 0.002 | 71 | 36 |
| sCD14 (ng/mL) | cpz_equivalent | 0 | 0 | -0.148 | 70.999 | 0.883 | 0.894 | 0 | 0 | 71 | 36 |

***Note*****:** cpz_equivalent = chlorpromazine equivalent dose

**Table S7.** Primary fixed effects linear mixed model, adjusting for duration of untreated psychosis (DUP) in the patient sample

| Biomarker | Term | estimate | Std error | statistic | df | p.value | p (FDR) | Conf. low | Conf. high | n | N subjects |
|---|---|---|---|---|---|---|---|---|---|---|---|
| BPI (ng/mL) | (Intercept) | 1.592 | 1.248 | 1.276 | 44.68 | 0.209 | 0.391 | -0.922 | 4.105 | 67 | 34 |
| BPI (ng/mL) | time2follow-upr | -0.052 | 0.126 | -0.415 | 46.5 | 0.68 | 0.763 | -0.306 | 0.201 | 67 | 34 |
| BPI (ng/mL) | age | 0.011 | 0.036 | 0.318 | 34.349 | 0.753 | 0.779 | -0.061 | 0.084 | 67 | 34 |
| BPI (ng/mL) | sexM | 0.534 | 0.568 | 0.94 | 33.625 | 0.354 | 0.508 | -0.621 | 1.689 | 67 | 34 |
| BPI (ng/mL) | bmi | 0.007 | 0.027 | 0.264 | 56.753 | 0.793 | 0.793 | -0.047 | 0.062 | 67 | 34 |
| BPI (ng/mL) | dup | 0.022 | 0.055 | 0.399 | 34.044 | 0.692 | 0.763 | -0.09 | 0.133 | 67 | 34 |
| Zonulin (ng/mL) | (Intercept) | 3.171 | 0.527 | 6.013 | 41.671 | **0** | **0.000** | 2.106 | 4.235 | 64 | 34 |
| Zonulin (ng/mL) | time2follow-up | 0.103 | 0.1 | 1.026 | 41.885 | 0.311 | 0.491 | -0.099 | 0.305 | 64 | 34 |
| Zonulin (ng/mL) | age | -0.018 | 0.013 | -1.36 | 34.013 | 0.183 | 0.391 | -0.045 | 0.009 | 64 | 34 |
| Zonulin (ng/mL) | sexM | -0.838 | 0.21 | -3.986 | 33.654 | 0 | **0.002** | -1.266 | -0.411 | 64 | 34 |
| Zonulin (ng/mL) | bmi | 0.108 | 0.016 | 6.736 | 55.91 | 0 | **0.000** | 0.076 | 0.14 | 64 | 34 |
| Zonulin (ng/mL) | dup | 0.015 | 0.02 | 0.714 | 33.972 | 0.48 | 0.629 | -0.027 | 0.056 | 64 | 34 |
| iFABP (ng/mL) | (Intercept) | -0.382 | 0.577 | -0.661 | 44.773 | 0.512 | 0.640 | -1.545 | 0.781 | 66 | 34 |
| iFABP (ng/mL) | time2follow-up | -0.206 | 0.063 | -3.273 | 45.384 | 0.002 | **0.010** | -0.333 | -0.079 | 66 | 34 |
| iFABP (ng/mL) | age | 0.022 | 0.016 | 1.364 | 33.34 | 0.182 | 0.391 | -0.011 | 0.055 | 66 | 34 |
| iFABP (ng/mL) | sexM | 0.73 | 0.257 | 2.845 | 32.775 | 0.008 | **0.033** | 0.208 | 1.252 | 66 | 34 |
| iFABP (ng/mL) | bmi | 0.013 | 0.013 | 0.931 | 60.528 | **0.356** | 0.508 | -0.014 | 0.04 | 66 | 34 |
| iFABP (ng/mL) | dup | 0.032 | 0.025 | 1.31 | 33.186 | 0.199 | 0.391 | -0.018 | 0.083 | 66 | 34 |

| | | | | | | | | | | | |
|---|---|---|---|---|---|---|---|---|---|---|---|
| LBP (ng/mL) | (Intercept) | 11.063 | 0.39 | 28.379 | 38.853 | 0 | 0.000 | 10.274 | 11.851 | 68 | 34 |
| LBP (ng/mL) | time2follow-up | 0.154 | 0.11 | 1.405 | 40.073 | 0.168 | 0.391 | -0.067 | 0.375 | 68 | 34 |
| LBP (ng/mL) | age | 0.004 | 0.009 | 0.372 | 34.171 | 0.712 | 0.763 | -0.016 | 0.023 | 68 | 34 |
| LBP (ng/mL) | sexM | -0.219 | 0.15 | -1.463 | 34.084 | 0.153 | 0.391 | -0.524 | 0.085 | 68 | 34 |
| LBP (ng/mL) | bmi | -0.007 | 0.012 | -0.595 | 43.01 | 0.555 | 0.666 | -0.032 | 0.018 | 68 | 34 |
| LBP (ng/mL) | dup | -0.01 | 0.015 | -0.711 | 34.336 | 0.482 | 0.629 | -0.04 | 0.019 | 68 | 34 |
| sCD14 (ng/mL) | (Intercept) | 7.681 | 0.078 | 99.009 | 38.38 | 0 | **0.000** | 7.524 | 7.838 | 68 | 34 |
| sCD14 (ng/mL) | time2follow-up | -0.031 | 0.026 | -1.226 | 38.525 | 0.228 | 0.402 | -0.083 | 0.02 | 68 | 34 |
| sCD14 (ng/mL) | age | -0.003 | 0.002 | -1.604 | 34.111 | 0.118 | 0.354 | -0.007 | 0.001 | 68 | 34 |
| sCD14 (ng/mL) | sexM | -0.078 | 0.03 | -2.636 | 34.047 | 0.013 | **0.047** | -0.138 | -0.018 | 68 | 34 |
| sCD14 (ng/mL) | bmi | -0.004 | 0.002 | -1.787 | 40.273 | 0.081 | 0.271 | -0.009 | 0.001 | 68 | 34 |
| sCD14 (ng/mL) | dup | 0.003 | 0.003 | 1.115 | 34.235 | 0.273 | 0.455 | -0.003 | 0.009 | 68 | 34 |

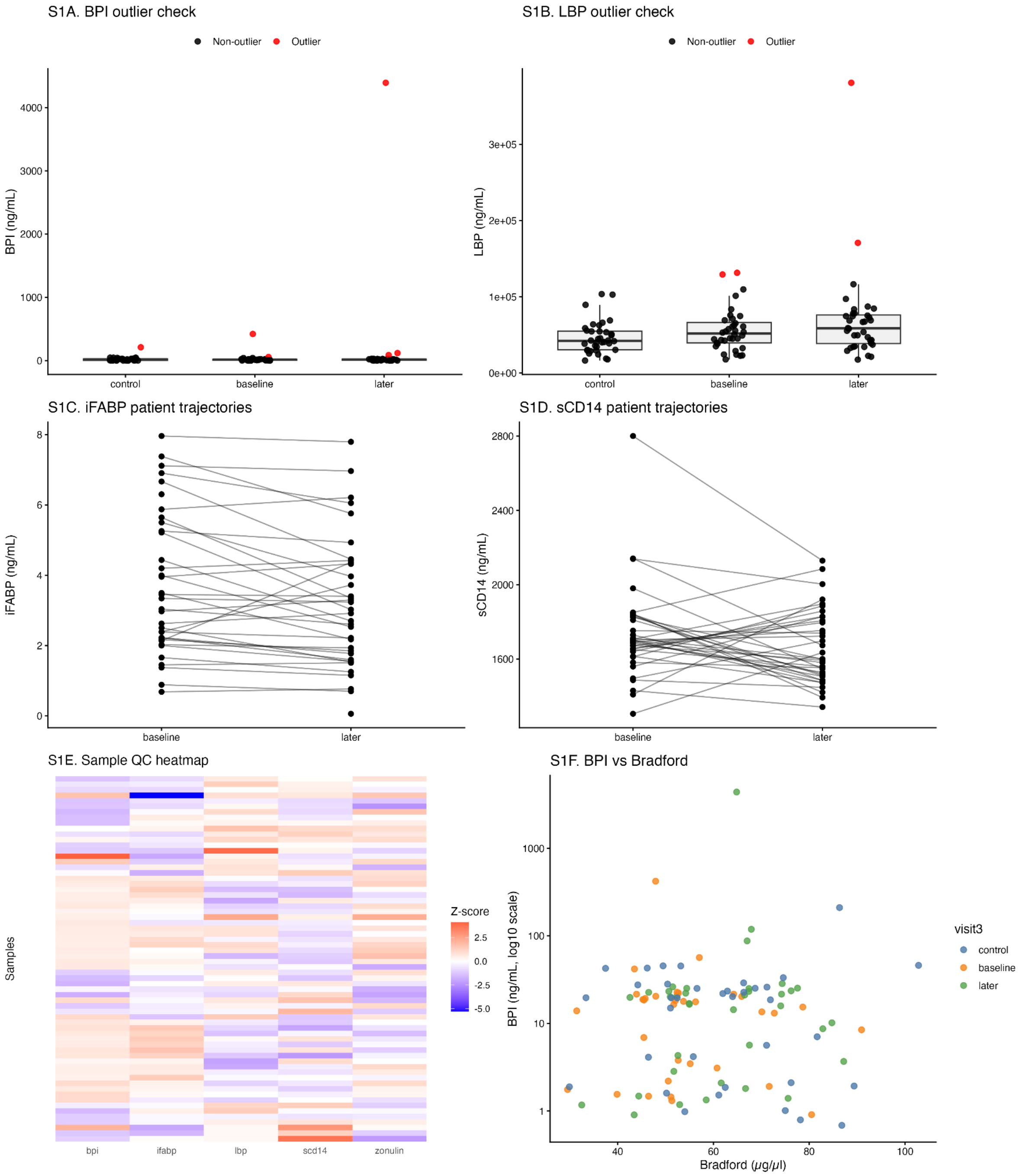


**Supplementary Figure S1. Quality control, outlier inspection, and sensitivity-related visualizations.**

**(A)** Outlier-highlighted distribution of BPI across controls, baseline patients, and follow-up patients. **(B)** Outlier-highlighted distribution of LBP across groups. **(C)** Within-patient longitudinal trajectories for iFABP. **(D)** Within-patient longitudinal trajectories for sCD14.

**(E)** Sample-level heatmap of standardized log-transformed biomarker values across all complete cases. **(F)** Relationship between BPI and Bradford protein concentration, shown to assess whether extreme biomarker values were explained by total protein content

**Table S8.** Spearman correlation coefficient (and FDR corrected p-value) between intestinal permeability indicators and clinical severity scales (only in the sample of patients, both time points)

| | GAF | PANSS positive | PANSS negative | PANSS Global | PANSS Total |
|---|---|---|---|---|---|
| BPI | -0.144 (0.550) | 0.211 (0.250) | -0.003 (0.992) | 0.147 (0.550) | 0.154 (0.542) |
| Zonulin | -0.057 (0.841) | -0.016 (0.955) | -0.098 (0.765) | -0.057 (0.841) | -0.043 (0.900) |
| iFABP | -0.187 (0.369) | 0.246 (0.148) | -0.001 (0.992) | 0.130 (0.626) | 0.121 (0.657) |
| LBP | -0.062 (0.841) | -0.115 (0.658) | 0.073 (0.825) | 0.029 (0.927) | 0.029 (0.927) |
| sCD14 | -0.015 (0.955) | 0.076 (0.841) | 0.063 (0.841) | 0.023 (0.952) | 0.061 (0.841) |

*Note.* Values represent Spearman's correlation coefficients (ρ), with FDR-adjusted p-values shown in parentheses. Intestinal permeability markers are reported in ng/mL. GAF = Global Assessment of Functioning scale; PANSS = Positive and Negative Syndrome Scale

**Table S9.** Spearman correlation coefficient (and FDR corrected p-value) between intestinal permeability markers and patient covariates (Age, BMI, duration of untreated psychosis, Bradford serum protein concentration)

| | **BPI** | **zonulin** | **iFABP** | **LBP** | **sCD14** | **NLR** | **age** | **BMI** | **DUP (months)** | **Bradford** | **Treatment duration (months)** |
|---|---|---|---|---|---|---|---|---|---|---|---|
| **BPI** | 1 (0) | 0.229 (0.238) | 0.162 (0.456) | -0.091 (0.673) | -0.134 (0.587) | 0.1 (0.673) | -0.067 (0.758) | 0.275 (0.093) | 0.148 (0.535) | 0.097 (0.673) | 0.113 (0.664) |
| **zonulin** | | 1 (0) | 0.053 (0.822) | 0.299 (0.077) | 0.017 (0.991) | -0.004 (0.991) | -0.006 (0.991) | **0.649 (0.000)** | 0.306 (0.077) | 0.129 (0.603) | 0.3 (0.084) |
| **iFABP** | | | 1 (0) | -0.315 (0.077) | -0.272 (0.096) | -0.172 (0.453) | 0.01 (0.991) | 0.301 (0.077) | 0.31 (0.077) | 0.055 (0.822) | -0.05 (0.822) |
| **LBP** | | | | 1 (0) | 0.262 (0.102) | 0.128 (0.603) | 0.119 (0.608) | -0.001 (0.991) | -0.13 (0.603) | -0.076 (0.717) | 0.105 (0.673) |
| **sCD14** | | | | | 1 (0) | 0.081 (0.716) | -0.093 (0.673) | -0.17 (0.451) | 0.088 (0.689) | 0.312 (0.077) | -0.174 (0.451) |
| **NLR** | | | | | | 1 (0) | -0.006 (0.991) | -0.093 (0.673) | -0.029 (0.936) | -0.214 (0.281) | -0.196 (0.380) |
| **age** | | | | | | | 1 (0) | 0.162 (0.456) | -0.077 (0.717) | -0.057 (0.815) | 0.101 (0.673) |
| **BMI** | | | | | | | | 1 (0) | 0.32 (0.077) | 0.029 (0.936) | 0.318 (0.077) |
| **DUP (months)** | | | | | | | | | 1 (0) | 0.337 (0.077) | -0.004 (0.991) |
| **Bradford** | | | | | | | | | | 1 (0) | 0.153 (0.524) |
| **Treatment duration (months)** | | | | | | | | | | | 1 (0) |

*Note:* Values represent Spearman's correlation coefficients (ρ), with FDR-adjusted p-values shown in parentheses. Intestinal permeability markers are reported in ng/mL; Bradford protein concentration is reported in µg/µL. Bold values indicate FDR-adjusted $p < 0.05$. NLR = neutrophil-to-lymphocyte ratio; DUP = duration of untreated psychosis.
